# Evaluating the causal association between type 2 diabetes and Alzheimer’s disease: a Two-Sample Mendelian Randomization Study

**DOI:** 10.1101/2024.12.19.24319309

**Authors:** Si Han, Tom Lelieveldt, Miriam Sturkenboom, Geert Jan Biessels, Fariba Ahmadizar

## Abstract

**Aims:** Type 2 diabetes mellitus (T2DM) and Alzheimer’s disease (AD) are significant global health issues. Epidemiological studies suggest T2DM increases AD risk, though confounding factors and reverse causality complicate this association. This study aims to clarify the causal relationship between T2DM and AD through a systematic review and meta-analysis of Mendelian randomization (MR) studies and a new two-sample MR analysis.

**Materials and Methods:** A literature search across major databases was conducted through May 2024 to identify MR studies linking T2DM and AD. Fixed/random-effect models provided pooled odds ratios (OR) with 95% confidence intervals (CI), and heterogeneity was assessed with the I² statistic. For our MR analysis, we pooled genetic variants from selected studies and analyzed AD outcomes using IGAP, EADB, and UKB databases. Multiple MR methods, including inverse-variance weighted (IVW) and pleiotropy-robust approaches, were applied for validation.

**Results:** Of 271 articles, eight MR studies were included (sample sizes: 68,905 to 788,989), all from European ancestry. Our meta-analysis found no significant causal link between T2DM and AD (OR = 1.01, 95% CI: 0.99-1.03) with moderate heterogeneity (I² = 44.16%). Similarly, our MR analysis using 511 SNPs as instrumental variables showed no significant associations in IGAP, EADB, or UKB data, consistent across sensitivity analyses.

**Conclusions:** This meta-MR and MR analysis revealed no significant causal association between T2DM and AD, indicating that T2DM may not directly influence AD risk. Further research should explore other mechanisms linking these conditions.

## Introduction

Type 2 diabetes mellitus (T2DM) is a chronic metabolic condition characterized by impaired insulin sensitivity and persistent hyperglycemia. As a significant public health challenge contributing to an increasing burden worldwide, the global diabetes prevalence in individuals aged 20-79 in 2021 was estimated to be 10.5% (536.6 million people), rising to 12.2% (783.2 million) in 2045 (1). In addition to its direct health impact and the increasing healthcare costs it brings, T2DM contributes to significant morbidity and mortality through its complications, such as cardiovascular disease, neuropathy, and kidney dysfunction, posing substantial challenges to healthcare systems and public health (1-3).

Alzheimer’s disease (AD), the most common form of dementia that contributes to 60-70% of dementia cases (4), is also a prevalent condition that significantly impacts global public health, marked by progressive cognitive decline and neurodegeneration (5).

Numerous epidemiological studies have reported the increased risk of dementia outcomes, especially AD, in individuals with T2DM, suggesting a potential link between these two conditions (6, 7). For instance, a meta-analysis reported a 73% higher risk of all types of dementia and a 56% increased risk of AD in individuals with T2DM (7, 8). This association has been observed consistently across diverse populations, pointing to T2DM as a significant risk factor for AD. Several biological mechanisms have been proposed to explain this relationship, including insulin resistance, chronic hyperglycemia, and inflammation (9-12).

Mendelian randomization (MR) analysis is a genetic epidemiology method that infers causal relationships between modifiable risk factors and health outcomes. By leveraging genetic variants as instrumental variables (IVs), MR minimizes confounding and reverse causation, common limitations in observational studies (13). While MR provides a robust method for inferring causality between T2DM and various complications, there remains a lack of comprehensive synthesis regarding its impact on cognitive outcomes, particularly AD (14-16). To address this gap, our study systematically reviewed and synthesized current research on the causal relationship between T2DM and AD by aggregating data from multiple published MR studies. One major limitation of meta-MR results is the risk of overestimating effect sizes due to the repeated inclusion of the same genetic variant, which can inflate associations and bias estimates (17, 18). To minimize this risk, we pooled single nucleotide polymorphisms (SNPs) associated with T2DM from MR studies included in this review and removed duplicate variants. Using the group of non-duplicate SNPs as IVs, we conducted a two-sample MR analysis to explore the unbiased causal association between T2DM and AD.

## Materials and Methods

### i) Systematic Review and Meta-MR

To identify all relevant articles that addressed the causal associations between exposure (T2DM) and outcome (AD), we systematically searched PubMed, Web of Science and the EMBASE regardless of language from inception until May 1, 2024 (the complete search strategy can be found in S1 Appendix). The reference list of MR studies included in this review was also searched manually for other potentially relevant inclusions. The Preferred Reporting Items for Systematic Reviews and Meta-Analyses (PRISMA) guidelines were followed. The registered ID in PROSPERO is CRD42024609885. The review protocal is available from the reuqest from author.

#### Search strategy

Two authors (SH and TL) independently implemented the search strategy. The process began with an initial screening of titles and abstracts, followed by an in-depth review of the full text for potential articles. Any disagreements between the two reviewers were addressed through discussion with a third author (FA), who provided adjudication.

#### Inclusion criteria

We included MR studies investigating the association between T2DM and AD. Eligible studies were required to report causal estimates, such as odds ratios (OR) or β-coefficients, presented as an absolute value per unit increase, along with the associated 95% confidence intervals (CI) or standard errors (SE). Only full original publications were considered for inclusion. In the case of duplicate cohorts, only the most recent MR studies with unique exposure-genome wide association studies (GWAS) and outcome-GWAS were retained for meta-MR.

#### Data Extraction

We extracted key details from each eligible MR study, including the first author, publication year, number of IVs, consortiums, sample size, population ancestry, MR design, analysis method, effect metrics (OR with 95% CI or β-coefficients with SEs), and sensitivity MR methods with their results.

#### Quality Assessment

We used the quality assessment tool incorporating ten questions designed to evaluate the quality of MR studies (19). Among the questions, three are core assumptions of MR: (1) the genetic variants used as IVs must be strongly associated with the exposure of interest, (2) the genetic variants should not be associated with confounding factors, and (3) the genetic variants should influence the outcome solely through their effect on the exposure. Studies failing to address these key assumptions were excluded from the analysis.

#### Statistical Analysis

The effect estimates were combined using either a fixed-effects or random-effects model depending on the heterogeneity among the included studies. Heterogeneity between studies was quantified using the I^2^ statistic with values greater than 75% representing high heterogeneity (20). For studies reporting β-coefficients and SEs, the ORs and their corresponding CIs were obtained by exponentiating the β-coefficients and their respective CIs.

### ii) Two-sample MR analysis

We extracted information from eight eligible MR studies included in our systematic review. This included SNPs, major and reference alleles, effect allele frequency, effect size, SEs, effect metrics, p-values, closest genes, chromosomes and locations, and sample sizes. One investigator (SH) extracted the data, which was verified by the third investigator (FA). Missing data were requested from corresponding authors via e-mail.

#### Instrumental variables selection

Based on three MR assumptions, we pooled genetic variants demonstrating genome-wide significant associations (p < 5×10^−8^) with T2DM (19, 21). These variants came from DIAbetes Genetics Replication And Meta-analysis (DIAGRAM), DIAbetes Meta-ANalysis of Trans-Ethnic association studies (DIAMANTE) consortia, and other studies (22-26). In these GWASs, various definitions of T2DM used across included studies, commonly based on diagnostic criteria such as fasting glucose (≥ 7.0 mmol/L), HbA1c (≥ 6.5%), or non-fasting glucose (≥ 11.1 mmol/L), or from medical records, hospital discharge data, and electronic health registries. Details of these consortiums can be found in the **S4 Appendix.** The study design of the current MR analysis can be found in **Fig 2**.

Given the overlapping of SNPs across the included studies, we employed a deduplication strategy based on the p-value associated with each SNP’s exposure (T2DM) and the corresponding GWAS sample size. In the final analysis, we retained only non-redundant SNPs, prioritizing those with the lowest p-values or derived from the most recent GWAS with the largest cohort size. 1,104 SNPs were initially merged from origin studies, with 859 proving to be unique. We implemented a standardisation process for the effect alleles to ensure comparability and accurate aggregation of genetic effect estimates across multiple cohorts.

To ensure the validity and robustness of the causal inference and to verify the assumptions underlying the MR approach, associated traits for the SNPs were manually verified using the GWAS catalog and PheWeb. These SNPs were then subjected to linkage disequilibrium (LD) clumping with a threshold of R^2^=0.1 and 1000 kilobases (kb). Following this clumping process, 511 SNPs remained. All SNPs demonstrated F-statistics greater than 10, indicating that the genetic variants explain a significant portion of the variance in the exposure variable. The details of IVs can be found in the **S5 Appendix**. In the sensitivity analysis, a broader range of R^2^ (from 0.01 to 0.8) was applied to capture a more comprehensive view of the association. By systematically varying the R^2^ thresholds, we aimed to balance the trade-off between instrument independence and coverage, thereby assessing the consistency of the causal estimates across varying degrees of SNP correlation.

#### Outcome Genetic Consortia Data

The International Genomics of Alzheimer’s Project (IGAP), European Alzheimer & Dementia Biobank (EADB) consortium and UK Biobank (UKB) from the included MR studies were utilized, which are all publicly available summary-level data (27-29). All the GWAS datasets used in this study obtained relevant ethics committee approvals, and participant informed consent at the time of their original data collection. IGAP is a comprehensive two-stage study based on GWASs of AD in individuals of European descent, which consists of Alzheimer Disease Genetics Consortium (ADGC), European Alzheimer’s Disease Initiative (EADI), and other consortiums (28, 30). In the first stage, IGAP utilized genotyped and imputed data on 7,055,881 SNPs to perform a meta-analysis of four previously published GWAS datasets, which included 17,008 AD cases and 37,154 controls.

EADB united various European cohorts and GWAS consortia, with summary estimates derived from 39,106 participants with clinically diagnosed AD, 46,828 participants with proxy AD, and 401,577 control participants without AD. Proxy AD was determined solely from the UKB through questionnaire data, where participants were asked if they had been diagnosed with AD or dementia.

UKB comprises 500,000 males and females from the general UK population, aged 40-69 at baseline (2006-2010). Cases were identified as algorithmically determined participants to have AD (N = 954), while non-cases were defined as participants who were not (N = 487,331). The analysis employed BOLT-LLM and was adjusted for age, sex, genotyping chip, and the top 10 genetic principal components, following the procedures of the Medical Research Council-Integrative Epidemiology Unit UK Biobank GWAS pipeline. Details about UKB and the pipeline can be found elsewhere (29, 31, 32).

#### Statistical Methods and Sensitivity Analyses

We harmonized the summary SNP-T2DM and SNP-AD statistics to ensure effect size alignment and prevent strand mismatch. In this analysis, we utilized proxy SNPs where the primary SNPs were unavailable in the outcome dataset, thereby enhancing SNP coverage and retaining relevant instruments that meet the LD threshold. A minor allele frequency (MAF) threshold of 0.01 was applied to ensure that SNPs with low allele frequencies, which could introduce noise or bias, were excluded from the analysis. In MR analysis, the inverse variance weighted (IVW) method was used as the primary analysis method. The IVW method operates under the assumption that all SNPs included in the causal estimate are valid instruments, implying that they do not violate any of the fundamental assumptions of MR.

To assess the potential impact of pleiotropy, we evaluated heterogeneity across SNP-specific MR estimates using Cochran’s Q statistic. We also performed MR-Egger regression, which provides a test for directional pleiotropy through its intercept, where a non-zero intercept suggests that pleiotropic effects are biasing the causal estimate (33, 34). We also used the weighted median estimator (WME), which allows up to 50% of the SNPs to be invalid instruments, offering a more robust causal estimate when pleiotropy is present (35, 36). The simple mode and weighted mode methods further complement this by assuming that the causal effect is determined by the most frequent estimate among the SNPs, with the weighted mode giving more importance to stronger instruments (37, 38).

All statistical analyses were performed using the “TwoSampleMR (0.5.10)” package in R Studio (version 2024.04). All P values were two-sided, and P < 0.05 was considered suggestive of statistical significance. The data, including exposures and outcomes, is all from open databases: https://www.ebi.ac.uk/gwas/home.

## Results

### i) Systematic Review and Meta-MR

The initial database search yielded 271 articles. Subsequent filtering to remove duplicates and articles not meeting the inclusion criteria. Further scrutiny for potential inclusions from reference lists led to 11 articles being considered for duplication cohorts’ check. Among these, one was excluded because of duplicate exposure and outcome consortium (Morris 2012 GWAS and IGAP); one employed the one-sample MR study design, and one didn’t use IVW as the primary analysis methods. Eight MR studies met all criteria and were selected for inclusion in the meta-MR and subsequent MR analysis (15, 32, 39-44). In these eight MR studies, total sample sizes, including case and control, range from 68,905 to 788,989, all of European ancestry. All the studies passed the quality assessment. Information on individual studies included in this review (consortium, sample size, IVs, study design, population, and main results) is shown in the **S2 Appendix**. The quality assessment questions and results are shown in the **S6 and S7 Appendix**. The PRISMA diagram is shown in **Fig 1**. The PRISMA checklist can be found in **S11 Appendix**. The estimates represent the OR of AD per 1-unit higher log odds of T2DM.

**Fig 1.**
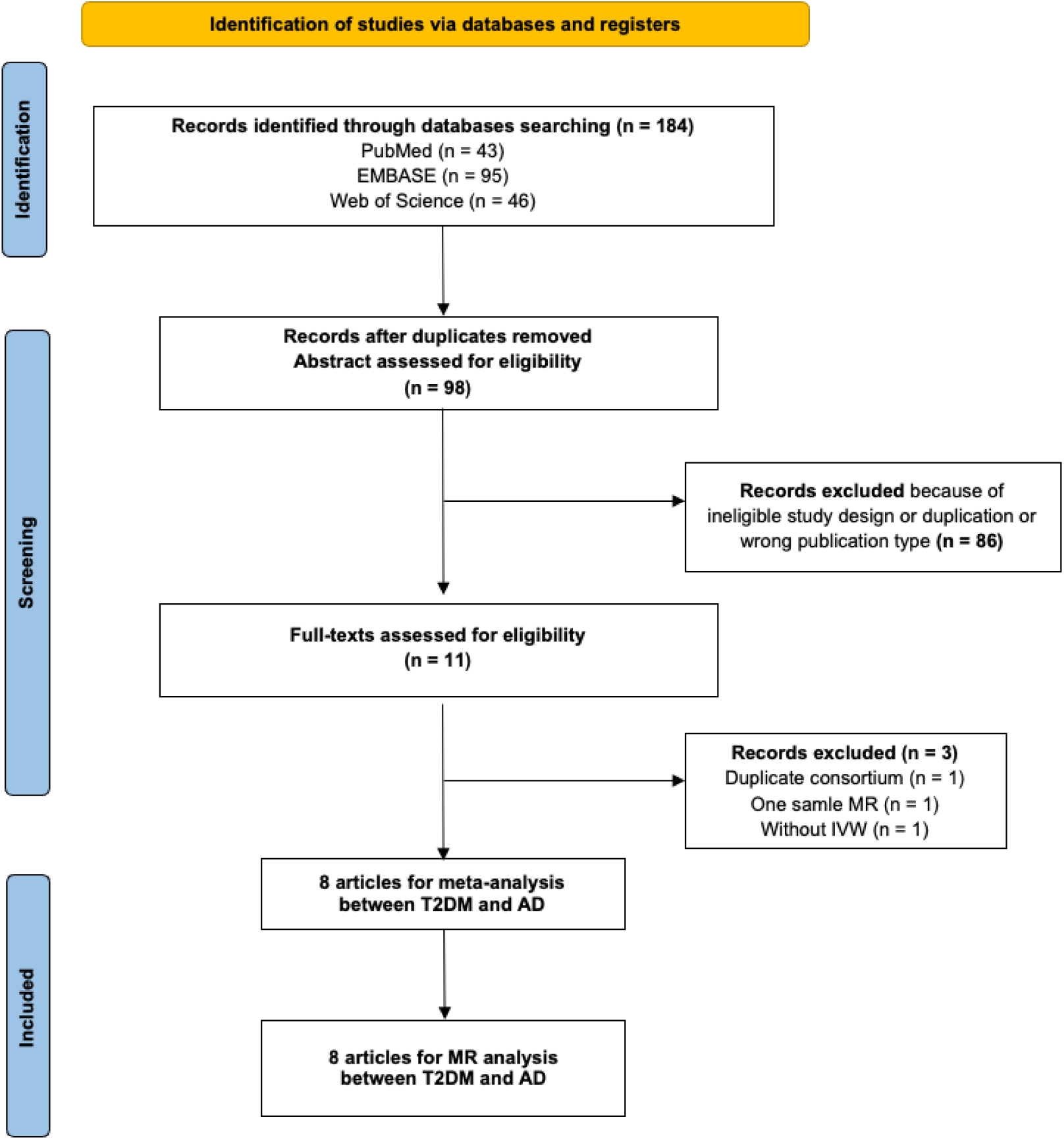
PRISMA ((Preferred Reporting Items for Systematic reviews and Meta-Analyses) flow diagram for systematic review and meta-analysis of the association between type 2 diabetes and Alzheimer’s disease. T2DM, type 2 diabetes mellitus; AD, Alzheimer’s disease; MR, mendelian randomization; IVW, inverse variance weighting.

**Fig 2.**
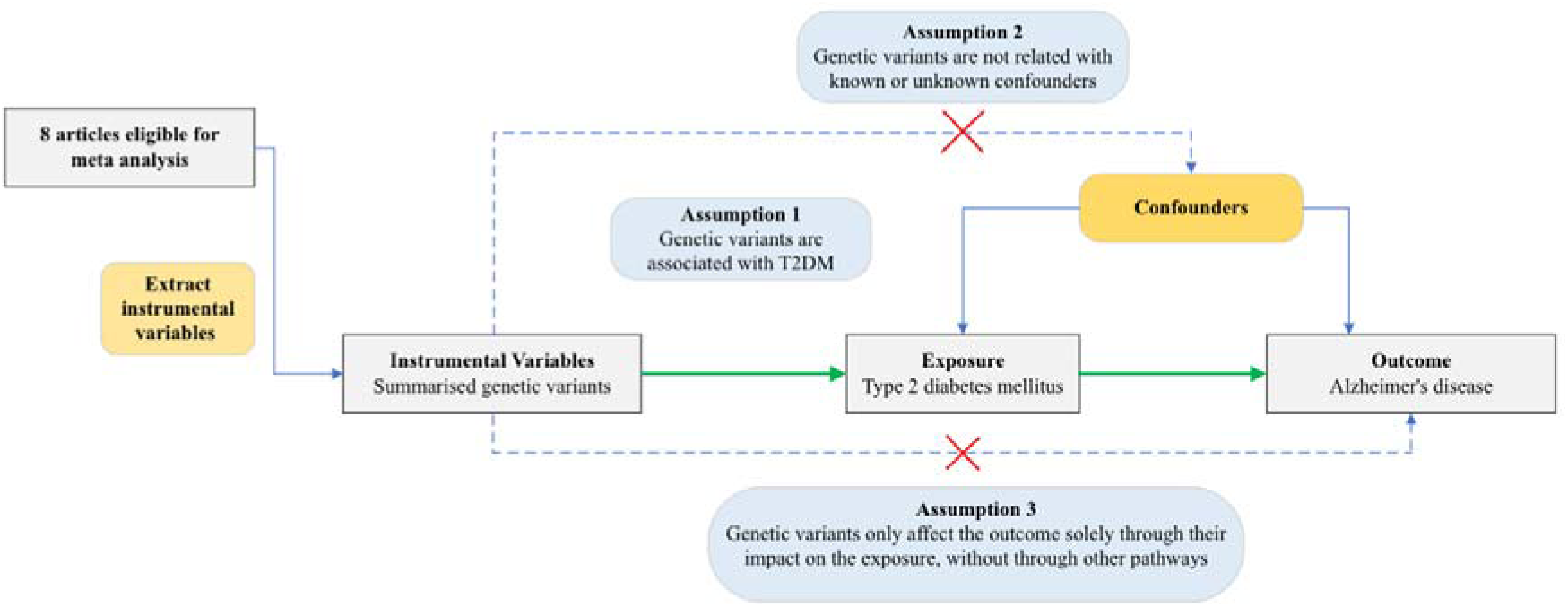
Flowchart of Mendelian Randomization analysis. T2DM, type 2 diabetes mellitus

In the meta-analysis, the Cochran’s Q test yielded a value of 10.74 with p-value 0.097, the I^2^ statistic was calculated to be 44.16%. Based on a fixed-effec model, the pooled risk estimate indicates that a genetic predisposition to T2DM was not significantly associated with an increased risk of AD (OR: 1.01; 95% CI: 0.99-1.03; p-value= 0.2) (**Fig 3**). The result of random effect model showed consistent results (**S3 Appendix**).

**Fig 3.**
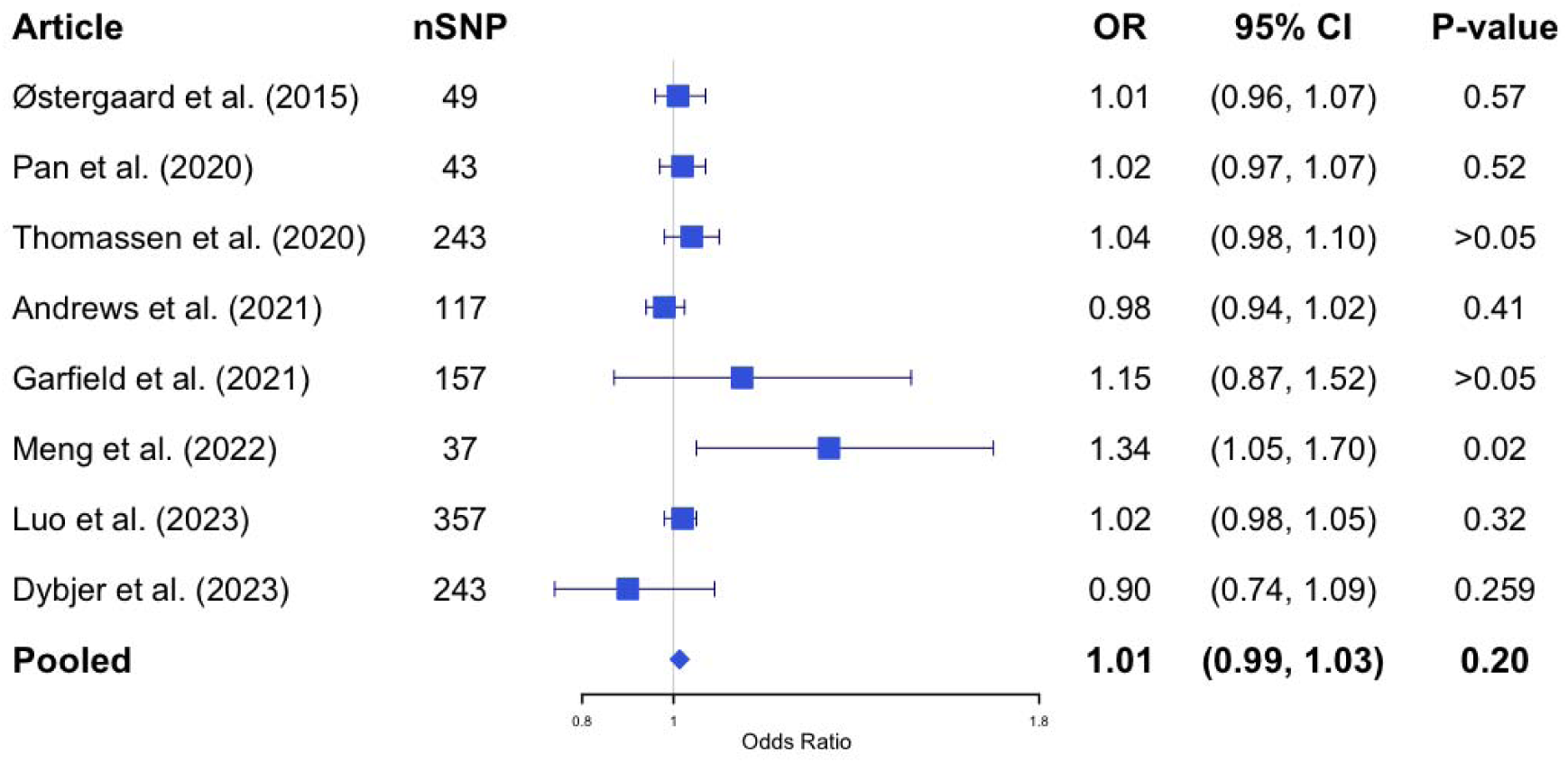
The pooled result of Mendelian Randomization analysis between type 2 diabetes mellitus and Alzheimer’s disease (based on fixed effect model with heterogeneity I^2^=44.16%) SNP, Single nucleotide polymorphism; OR, odds ratio; CI, confidence interval

### ii) Two-Sample MR analysis

IGAP dataset: The results of our MR study using the IGAP dataset, which included 440 SNPs as IVs, are presented in the **S8 Appendix** and visualized in **Fig 4a**. Although 511 SNPs were initially identified as IVs after clumping (R^2^ =0.1), the final analysis included fewer SNPs due to several factors. First, not all SNPs from the clumped list had corresponding outcome data in the outcome dataset, leading to the exclusion of SNPs without matching outcome information. Additionally, despite choosing proxies where possible, some SNPs lacked suitable proxies with sufficient LD, resulting in further reduction.

**Fig 4.**
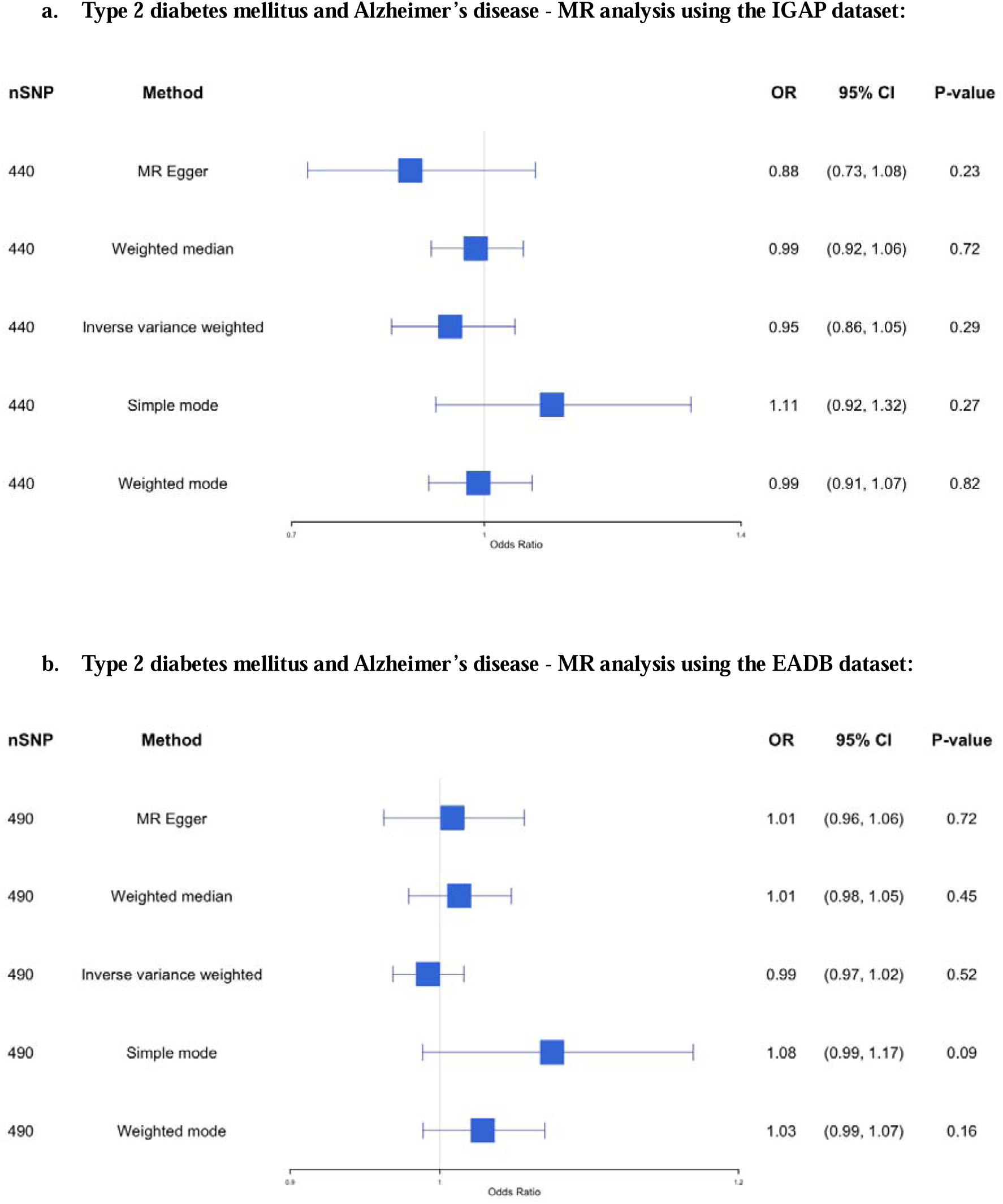
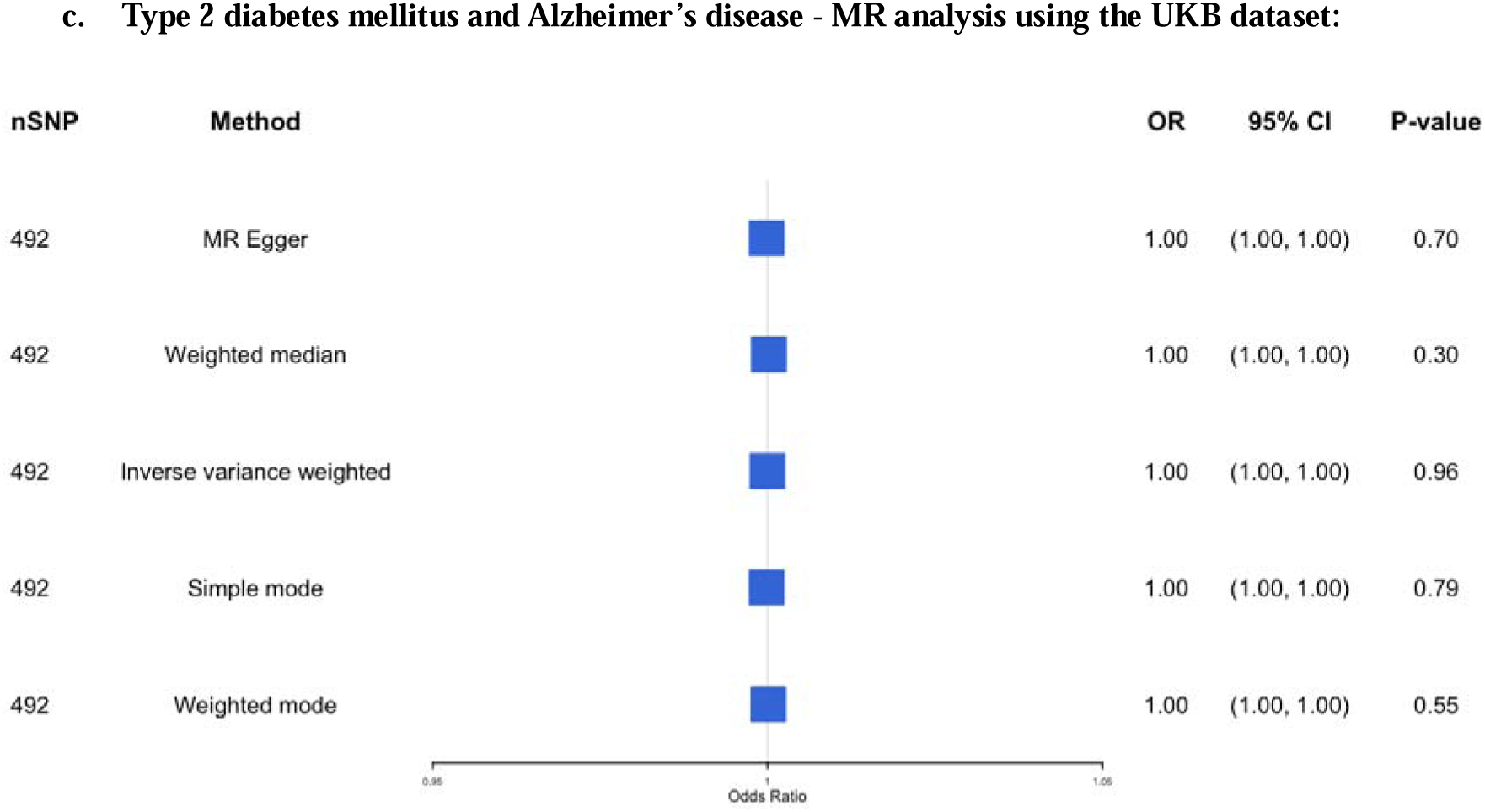
The causal association between type 2 diabetes mellitus and Alzheimer’s disease - MR analysis using the IGAP dataset, EADB dataset, and UKB dataset. SNP, Single nucleotide polymorphism; OR, odds ratio; CI, confidence interval; IGAP, The International Genomics of Alzheimer’s Project; EADB, European Alzheimer & Dementia Biobank consortium; UKB, UK Biobank

Our findings revealed no significant causal association (OR 0.95; 95% CI: 0.86-1.05; p-value = 0.29) between genetic predisposition to T2DM and AD using the IVW method, even after applying multiple MR methods to assess the robustness of the results. The result remains insignificant in the sensitivity analysis with different clumping R^2^ (ranging from 0.01 to 0.8). The details of the results can be found in **Fig 5a-5e**.

**Fig 5.**
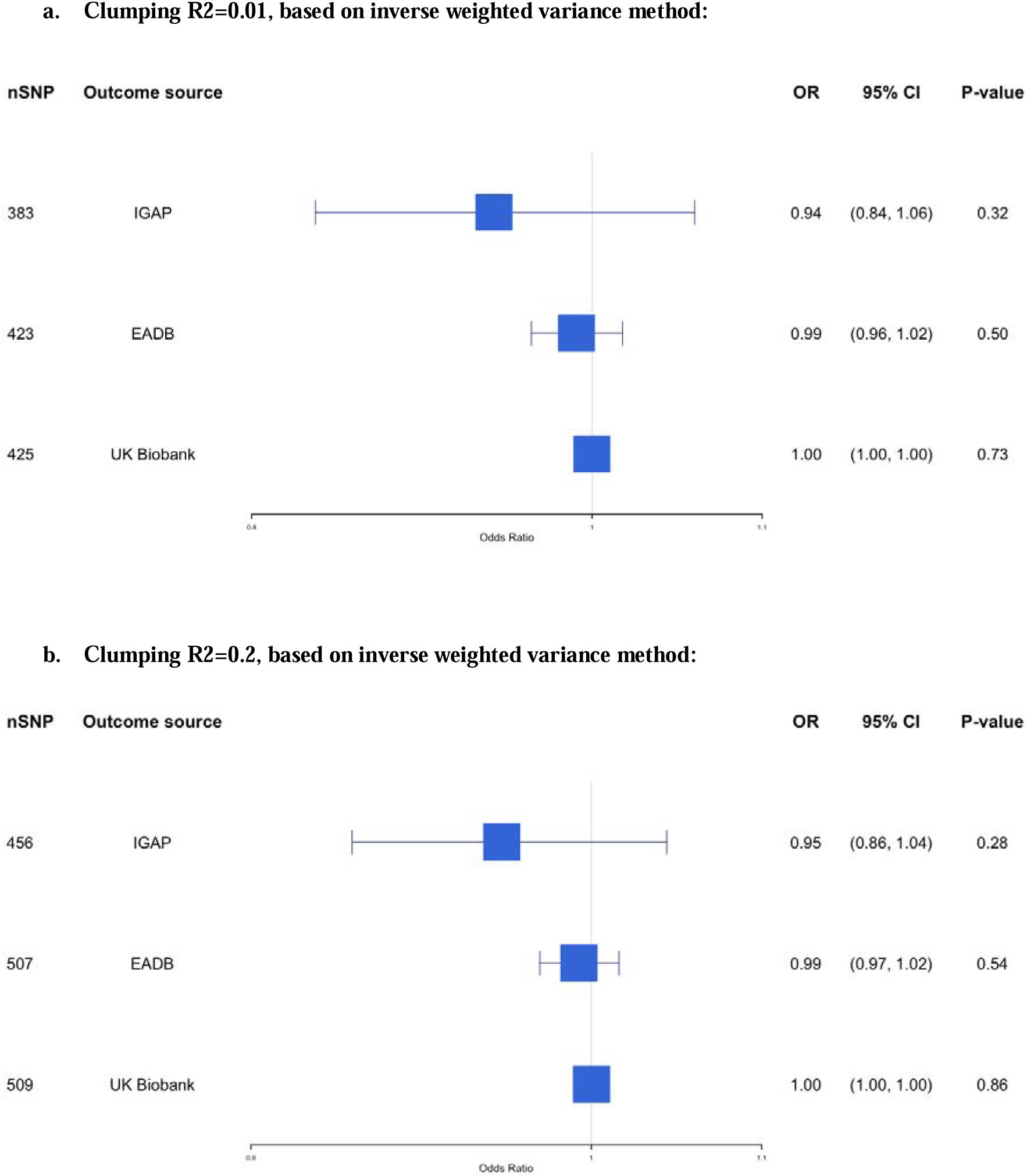
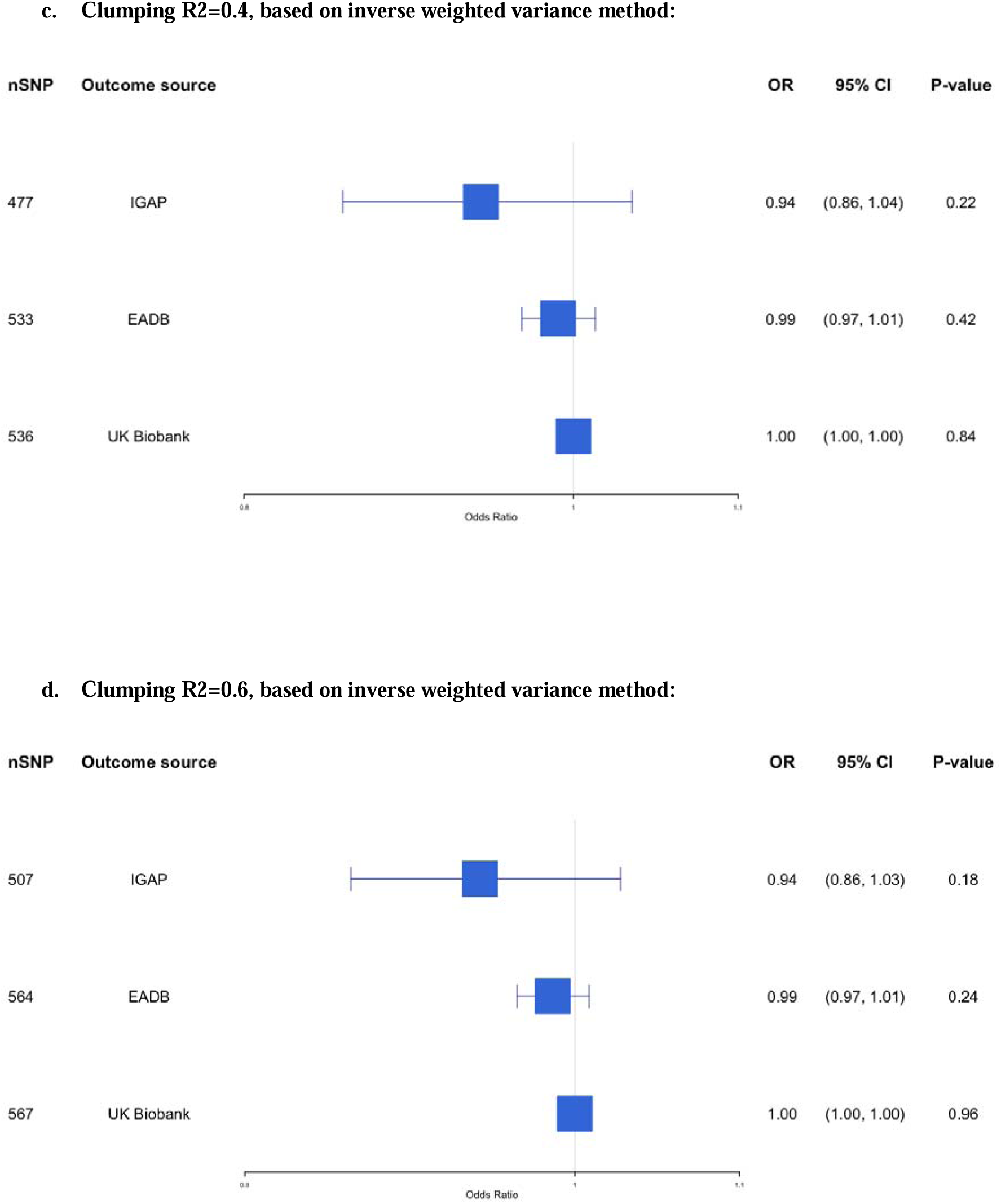
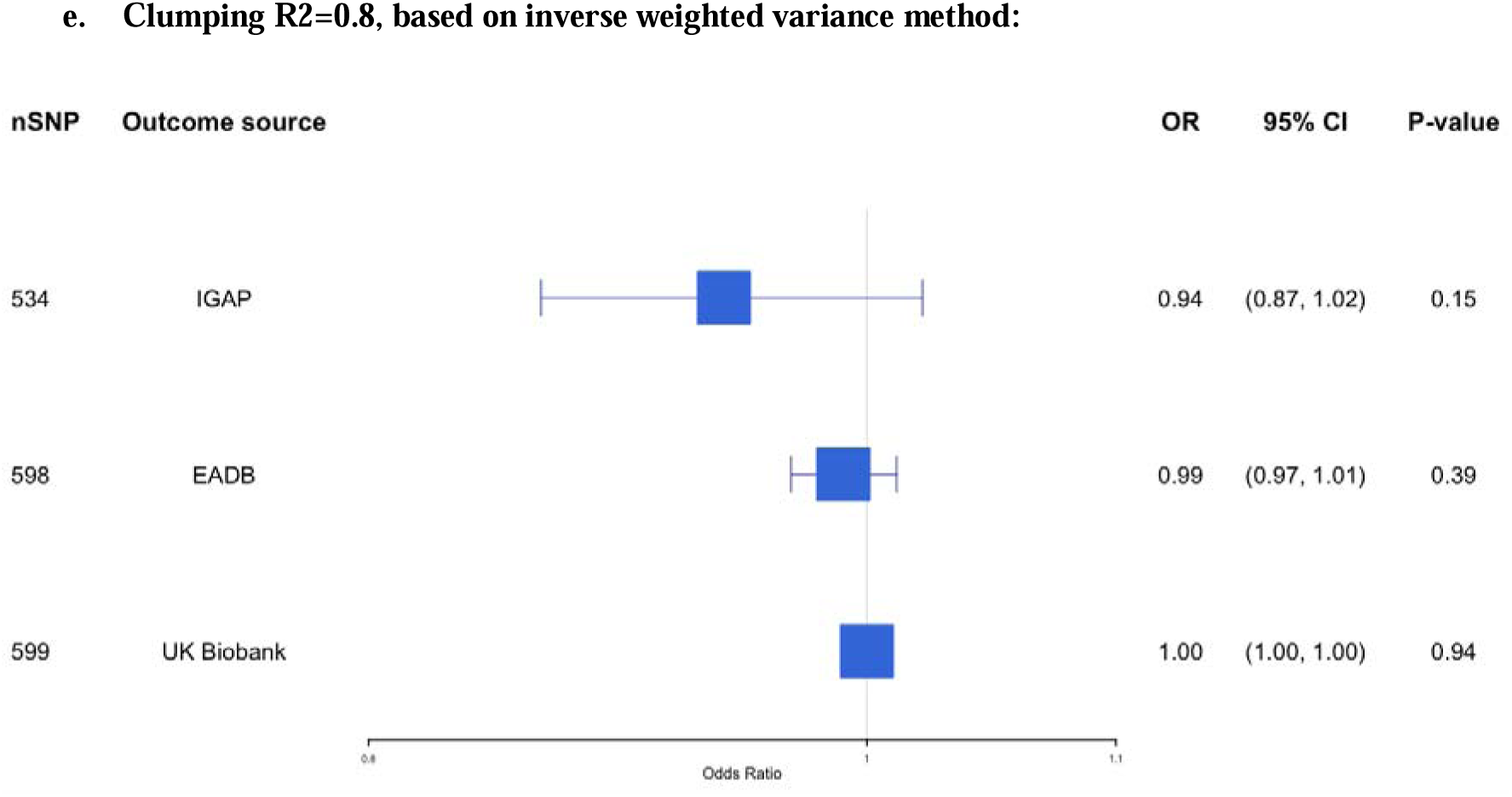
The causal association between type 2 diabetes mellitus and Alzheimer’s disease (clumping R2=0.01, 0.2, 0.4, 0.6, and 0.8, based on inverse weighted variance method). SNP, Single nucleotide polymorphism; OR, odds ratio; CI, confidence interval; IGAP, The International Genomics of Alzheimer’s Project; EADB, European Alzheimer & Dementia Biobank consortium

EADB dataset: The results of this MR study, which included 490 SNPs as IVs, are presented in the **S9 Appendix** and visualized in **Fig 4b**. No significant causal association was found using the IVW method (OR 0.99; 95% CI 0.97-1.02; p-value= 0.52) also across all MR methods. The result remains insignificant in the sensitivity analysis with different clumping R^2^ (ranging from 0.01 to 0.8). The details of the results can be found in **Fig 5a-5e**.

UKB dataset: The results from the UKB dataset, which included 492 SNPs as IVs, are presented in the **S10 Appendix** and visualized in **Fig 4c**, showing no significant association (OR 1.00; 95% CI 1.00-1.00; p-value = 0.96). The result remains insignificant in the sensitivity analysis with different clumping R^2^ (ranging from 0.01 to 0.8). The details of the results can be found in **Fig 5a-5e**.

No significant pleiotropy was observed in the above analyses, with the p-value of MR Egger regression above 0.05.

## Discussion

Due to the nature of MR, which leverages genetic variants as proxies for exposures, this method can help address potential causal relationships between risk factors (T2DM) and outcomes (AD), minimizing the influence of confounding and reverse causality. Our study provides a thorough evaluation of the potential causal association between T2DM and AD by presenting findings of a meta-MR, as well as a new two-sample MR analysis based on IVs for T2DM identified from our review and outcome data from three large datasets (IGAP, EADB, UKB). In the meta-analysis of eight MR studies, we did not observe a statistically significant causal association between genetic predisposition to T2DM and AD. Similarly, our two-sample MR analyses revealed no statistically significant support for a causal association between T2DM and AD across various MR methods, such as IVW, MR-Egger, and weighted median. The results remained consistent even after employing different clumping thresholds (R² ranging from 0.01 to 0.8), further suggesting that the genetic predisposition to T2DM does not have a strong causal impact on AD development. Notably, no evidence of directional pleiotropy, as indicated by non-significant MR-Egger intercepts across all datasets, enhancing our findings’ reliability.

With these results, it is essential to assess the accuracy of our findings and investigate reasons for discrepancies with previous MR studies; one of the included MR studies in our review obtained significant association, which is opposite to other included studies (41). Firstly, the choice of genetic variants used as IVs differs between studies. Genetic variants with varying strengths of association with T2DM could lead to inconsistent results, particularly if some studies utilize weak IVs that violate MR assumptions. Secondly, variations in data quality and population characteristics such as ancestry, age, and sex could also contribute to differing findings. Studies with more homogenous populations may yield stronger associations than those with diverse cohorts, where confounding factors might obscure true relationships. Our review showed that studies with different outcomes-GWAS may yield wider CIs (32). Thirdly, the definition of AD can affect the result of the MR analysis. Among the MR studies included in this paper, some studies used proxy AD diagnosis, which may affect the characteristics of the population (15).

While our study did not find strong evidence of a direct causal association, epidemiological studies frequently report links between T2DM and AD. These associations are likely driven by residual confounding factors such as age, obesity, and hypertension, which can contribute to common underlying mechanisms like metabolic dysfunction, inflammation, and vascular damage (6, 45). One possible explanation for the lack of a significant causal link could be that T2DM may influence dementia risk through pathways distinct from those involved in AD, including insulin resistance (46-48), chronic hyperglycemia (49, 50), inflammation (51-53), and vascular dysfunction (54-57). T2DM is mainly associated with vascular changes that elevate the risk for vascular dementia rather than AD. Conditions like vascular injury and small vessel disease, prevalent in individuals with T2DM, may contribute to cognitive decline and dementia through cerebrovascular damage rather than through amyloid or tau pathology, which is central to AD. Studies indicate that while the pathological changes observed in AD and vascular dementia can coexist, their mechanistic pathways may diverge, which may explain why our MR analysis did not find an increased risk of AD associated with T2DM.

Genetic instruments in MR studies may capture broader metabolic traits linked with T2DM, such as insulin resistance, obesity, and dyslipidemia, which could have complex effects on brain health. While these metabolic dysfunctions may increase overall dementia risk, their impact on AD pathology might be indirect or less significant. Additionally, T2DM is influenced by genetic, lifestyle, and environmental factors, and genetic variants linked to T2DM may not fully reflect the molecular mechanisms underlying AD, potentially diluting any AD-specific effect.

### Clinical Relevance

The lack of a significant causal association between T2DM and AD in our study suggests that T2DM may not directly contribute to AD development. This finding challenges the common assumption that diabetes is a direct risk factor for AD and necessitates a reevaluation of the implications of T2DM for cognitive health. However, it is crucial to note that our results do not rule out the possibility that T2DM could increase dementia risk through alternative pathways.

Potential mechanisms, including insulin resistance, chronic hyperglycemia, vascular damage, and inflammation, may independently contribute to cognitive decline, offering an explanation for the associations observed in epidemiological studies between T2DM and dementia risk. While our findings do not support a direct link between T2DM and AD, they underscore the need for further research into the broader impact of T2DM on dementia, particularly through non-AD pathways (58-60). Future studies should focus on the differential effects of T2DM on various dementia types to better understand its role in cognitive decline, which could provide valuable insights for healthcare providers and inform strategies for prevention and intervention in at-risk populations.

### Strengths and Limitations

A key strength of our study is its comprehensive methodology, which combines a systematic review and meta-analysis of existing MR studies to enhance statistical power and produce robust association estimates. Our original MR analyses conducted across multiple large-scale datasets further strengthen this. Our rigorous sensitivity analyses, utilizing various MR methods and clumping thresholds, also help mitigate potential biases such as horizontal pleiotropy. However, the limitation should be noted. First, moderate heterogeneity among MR studies was observed in our meta-MR study. We conducted additional sensitivity analysis using a random-effects model, which produced consistent results, supporting the robustness of the pooled effect estimate. Second, the generalizability of our findings may be restricted, particularly for populations outside of European ancestry, necessitating caution in applying these results broadly.

### Conclusions

In conclusion, no causal association was observed in our study, which included a meta-MR and a two-sample MR analysis of T2DM and AD using large, well-powered datasets. These findings highlight the need for further research to explore other potential mechanisms linking metabolic disorders and neurodegenerative diseases, such as shared inflammatory or vascular pathways, rather than direct genetic predispositions to T2DM as a cause of AD.

## Data Availability

All data produced in the present study are available upon reasonable request to the authors

## Author Contributions

F.A. conceived of the presented idea. S.H. and T.L. did the systematic review. S.H. conducted all statistical analyses under the supervision of F.A. and G.B in close collaboration with a genetic epidemiologist as a consultant. All authors discussed the results and contributed to the final manuscript.

## References

1. Sun H, Saeedi P, Karuranga S, Pinkepank M, Ogurtsova K, Duncan BB, et al. IDF Diabetes Atlas: Global, regional and country-level diabetes prevalence estimates for 2021 and projections for 2045. Diabetes Res Clin Pract. 2022;183:109119.

2. Global, regional, and national burden of diabetes from 1990 to 2021, with projections of prevalence to 2050: a systematic analysis for the Global Burden of Disease Study 2021. Lancet. 2023;402(10397):203-34.

3. Zakir M, Ahuja N, Surksha MA, Sachdev R, Kalariya Y, Nasir M, et al. Cardiovascular Complications of Diabetes: From Microvascular to Macrovascular Pathways. Cureus. 2023;15(9):e45835.

4. (WHO) WHO. Dementia 2019 [Available from: https://www.who.int/news-room/fact-sheets/detail/dementia. [

5. Kumar A SJ, Lui F, et al. Alzheimer Disease. [Updated 2024 Feb 12]. In: StatPearls [Internet]. Treasure Island (FL): StatPearls Publishing; 2024 Jan-. Available from: https://www.ncbi.nlm.nih.gov/books/NBK499922/.

6. Rojas M, Chávez-Castillo M, Bautista J, Ortega Á, Nava M, Salazar J, et al. Alzheimer’s disease and type 2 diabetes mellitus: Pathophysiologic and pharmacotherapeutics links. World J Diabetes. 2021;12(6):745–66.

7. Gudala K, Bansal D, Schifano F, Bhansali A. Diabetes mellitus and risk of dementia: A meta-analysis of prospective observational studies. J Diabetes Investig. 2013;4(6):640–50.

8. Biessels GJ, Staekenborg S, Brunner E, Brayne C, Scheltens P. Risk of dementia in diabetes mellitus: a systematic review. Lancet Neurol. 2006;5(1):64–74.

9. Jayaraman A, Pike CJ. Alzheimer’s disease and type 2 diabetes: multiple mechanisms contribute to interactions. Curr Diab Rep. 2014;14(4):476.

10. Umegaki H. Neurodegeneration in diabetes mellitus. Adv Exp Med Biol. 2012;724:258–65.

11. Singh VP, Bali A, Singh N, Jaggi AS. Advanced glycation end products and diabetic complications. Korean J Physiol Pharmacol. 2014;18(1):1–14.

12. Biessels GJ, Despa F. Cognitive decline and dementia in diabetes mellitus: mechanisms and clinical implications. Nat Rev Endocrinol. 2018;14(10):591–604.

13. Sanderson E, Glymour MM, Holmes MV, Kang H, Morrison J, Munafò MR, et al. Mendelian randomization. Nat Rev Methods Primers. 2022;2.

14. Litkowski EM, Logue MW, Zhang R, Charest BR, Lange EM, Hokanson JE, et al. Mendelian randomization study of diabetes and dementia in the Million Veteran Program. Alzheimer’s & Dementia. 2023;19(10):4367–76.

15. Luo J, Thomassen JQ, Bellenguez C, Grenier-Boley B, de Rojas I, Castillo A, et al. Genetic Associations Between Modifiable Risk Factors and Alzheimer Disease. JAMA Netw Open. 2023;6(5):e2313734.

16. Hardy J, de Strooper B, Escott-Price V. Diabetes and Alzheimer’s disease: shared genetic susceptibility? Lancet Neurol. 2022;21(11):962–4.

17. Stringer S, Wray NR, Kahn RS, Derks EM. Underestimated effect sizes in GWAS: fundamental limitations of single SNP analysis for dichotomous phenotypes. PLoS One. 2011;6(11):e27964.

18. Uffelmann E, Huang QQ, Munung NS, de Vries J, Okada Y, Martin AR, et al. Genome-wide association studies. Nature Reviews Methods Primers. 2021;1(1):59.

19. Davies NM, Holmes MV, Davey Smith G. Reading Mendelian randomisation studies: a guide, glossary, and checklist for clinicians. Bmj. 2018;362:k601.

20. Higgins JP, Thompson SG, Deeks JJ, Altman DG. Measuring inconsistency in meta-analyses. Bmj. 2003;327(7414):557-60.

21. Burgess S, Davey Smith G, Davies NM, Dudbridge F, Gill D, Glymour MM, et al. Guidelines for performing Mendelian randomization investigations: update for summer 2023. Wellcome Open Res. 2019;4:186.

22. Morris AP, Voight BF, Teslovich TM, Ferreira T, Segrè AV, Steinthorsdottir V, et al. Large-scale association analysis provides insights into the genetic architecture and pathophysiology of type 2 diabetes. Nat Genet. 2012;44(9):981–90.

23. Scott RA, Scott LJ, Mägi R, Marullo L, Gaulton KJ, Kaakinen M, et al. An Expanded Genome-Wide Association Study of Type 2 Diabetes in Europeans. Diabetes. 2017;66(11):2888–902.

24. Mahajan A, Wessel J, Willems SM, Zhao W, Robertson NR, Chu AY, et al. Refining the accuracy of validated target identification through coding variant fine-mapping in type 2 diabetes. Nat Genet. 2018;50(4):559–71.

25. Xue A, Wu Y, Zhu Z, Zhang F, Kemper KE, Zheng Z, et al. Genome-wide association analyses identify 143 risk variants and putative regulatory mechanisms for type 2 diabetes. Nat Commun. 2018;9(1):2941.

26. Vujkovic M, Keaton JM, Lynch JA, Miller DR, Zhou J, Tcheandjieu C, et al. Discovery of 318 new risk loci for type 2 diabetes and related vascular outcomes among 1.4 million participants in a multi-ancestry meta-analysis. Nat Genet. 2020;52(7):680–91.

27. Bellenguez C, Küçükali F, Jansen IE, Kleineidam L, Moreno-Grau S, Amin N, et al. New insights into the genetic etiology of Alzheimer’s disease and related dementias. Nat Genet. 2022;54(4):412–36.

28. Kunkle BW, Grenier-Boley B, Sims R, Bis JC, Damotte V, Naj AC, et al. Genetic meta-analysis of diagnosed Alzheimer’s disease identifies new risk loci and implicates Aβ, tau, immunity and lipid processing. Nat Genet. 2019;51(3):414–30.

29. Larsson SC, Woolf B, Gill D. Plasma Caffeine Levels and Risk of Alzheimer’s Disease and Parkinson’s Disease: Mendelian Randomization Study. Nutrients. 2022;14(9).

30. Lambert JC, Ibrahim-Verbaas CA, Harold D, Naj AC, Sims R, Bellenguez C, et al. Meta-analysis of 74,046 individuals identifies 11 new susceptibility loci for Alzheimer’s disease. Nat Genet. 2013;45(12):1452–8.

31. Bycroft C, Freeman C, Petkova D, Band G, Elliott LT, Sharp K, et al. The UK Biobank resource with deep phenotyping and genomic data. Nature. 2018;562(7726):203-9.

32. Garfield V, Farmaki AE, Fatemifar G, Eastwood SV, Mathur R, Rentsch CT, et al. Relationship Between Glycemia and Cognitive Function, Structural Brain Outcomes, and Dementia: A Mendelian Randomization Study in the UK Biobank. Diabetes. 2021;70(10):2313–21.

33. Schmidt AF, Dudbridge F. Mendelian randomization with Egger pleiotropy correction and weakly informative Bayesian priors. Int J Epidemiol. 2018;47(4):1217–28.

34. Hemani G, Bowden J, Davey Smith G. Evaluating the potential role of pleiotropy in Mendelian randomization studies. Hum Mol Genet. 2018;27(R2):R195–r208.

35. Bowden J, Davey Smith G, Burgess S. Mendelian randomization with invalid instruments: effect estimation and bias detection through Egger regression. Int J Epidemiol. 2015;44(2):512–25.

36. Slob EAW, Burgess S. A comparison of robust Mendelian randomization methods using summary data. Genet Epidemiol. 2020;44(4):313–29.

37. Hartwig FP, Davey Smith G, Bowden J. Robust inference in summary data Mendelian randomization via the zero modal pleiotropy assumption. Int J Epidemiol. 2017;46(6):1985–98.

38. Boehm FJ, Zhou X. Statistical methods for Mendelian randomization in genome-wide association studies: A review. Comput Struct Biotechnol J. 2022;20:2338–51.

39. Østergaard SD, Mukherjee S, Sharp SJ, Proitsi P, Lotta LA, Day F, et al. Associations between Potentially Modifiable Risk Factors and Alzheimer Disease: A Mendelian Randomization Study. PLoS Med. 2015;12(6):e1001841; discussion e.

40. Andrews SJ, Fulton-Howard B, O’Reilly P, Marcora E, Goate AM. Causal Associations Between Modifiable Risk Factors and the Alzheimer’s Phenome. Ann Neurol. 2021;89(1):54–65.

41. Meng L, Wang Z, Ji HF, Shen L. Causal association evaluation of diabetes with Alzheimer’s disease and genetic analysis of antidiabetic drugs against Alzheimer’s disease. Cell Biosci. 2022;12(1):28.

42. Pan Y, Chen W, Yan H, Wang M, Xiang X. Glycemic traits and Alzheimer’s disease: a Mendelian randomization study. Aging (Albany NY). 2020;12(22):22688–99.

43. Dybjer E, Kumar A, Nägga K, Engström G, Mattsson-Carlgren N, Nilsson PM, et al. Polygenic risk of type 2 diabetes is associated with incident vascular dementia: a prospective cohort study. Brain Commun. 2023;5(2):fcad054.

44. Thomassen JQ, Tolstrup JS, Benn M, Frikke-Schmidt R. Type-2 diabetes and risk of dementia: observational and Mendelian randomisation studies in 1 million individuals. Epidemiol Psychiatr Sci. 2020;29:e118.

45. Wang F, Guo X, Shen X, Kream RM, Mantione KJ, Stefano GB. Vascular dysfunction associated with type 2 diabetes and Alzheimer’s disease: a potential etiological linkage. Med Sci Monit Basic Res. 2014;20:118–29.

46. Milstein JL, Ferris HA. The brain as an insulin-sensitive metabolic organ. Mol Metab. 2021;52:101234.

47. Barone E, Di Domenico F, Perluigi M, Butterfield DA. The interplay among oxidative stress, brain insulin resistance and AMPK dysfunction contribute to neurodegeneration in type 2 diabetes and Alzheimer disease. Free Radic Biol Med. 2021;176:16–33.

48. Berlanga-Acosta J, Guillén-Nieto G, Rodríguez-Rodríguez N, Bringas-Vega ML, García-Del-Barco-Herrera D, Berlanga-Saez JO, et al. Insulin Resistance at the Crossroad of Alzheimer Disease Pathology: A Review. Front Endocrinol (Lausanne). 2020;11:560375.

49. González P, Lozano P, Ros G, Solano F. Hyperglycemia and Oxidative Stress: An Integral, Updated and Critical Overview of Their Metabolic Interconnections. Int J Mol Sci. 2023;24(11).

50. Caturano A, D’Angelo M, Mormone A, Russo V, Mollica MP, Salvatore T, et al. Oxidative Stress in Type 2 Diabetes: Impacts from Pathogenesis to Lifestyle Modifications. Curr Issues Mol Biol. 2023;45(8):6651–66.

51. Okdahl T, Wegeberg AM, Pociot F, Brock B, Størling J, Brock C. Low-grade inflammation in type 2 diabetes: a cross-sectional study from a Danish diabetes outpatient clinic. BMJ Open. 2022;12(12):e062188.

52. Tsalamandris S, Antonopoulos AS, Oikonomou E, Papamikroulis GA, Vogiatzi G, Papaioannou S, et al. The Role of Inflammation in Diabetes: Current Concepts and Future Perspectives. Eur Cardiol. 2019;14(1):50–9.

53. Rohm TV, Meier DT, Olefsky JM, Donath MY. Inflammation in obesity, diabetes, and related disorders. Immunity. 2022;55(1):31–55.

54. Albai O, Frandes M, Timar R, Roman D, Timar B. Risk factors for developing dementia in type 2 diabetes mellitus patients with mild cognitive impairment. Neuropsychiatr Dis Treat. 2019;15:167–75.

55. Ortiz GG, Huerta M, González-Usigli HA, Torres-Sánchez ED, Delgado-Lara DL, Pacheco-Moisés FP, et al. Cognitive disorder and dementia in type 2 diabetes mellitus. World J Diabetes. 2022;13(4):319–37.

56. Govindpani K, McNamara LG, Smith NR, Vinnakota C, Waldvogel HJ, Faull RL, et al. Vascular Dysfunction in Alzheimer’s Disease: A Prelude to the Pathological Process or a Consequence of It? J Clin Med. 2019;8(5).

57. Eisenmenger LB, Peret A, Famakin BM, Spahic A, Roberts GS, Bockholt JH, et al. Vascular contributions to Alzheimer’s disease. Transl Res. 2023;254:41–53.

58. Li Y, Liu Y, Liu S, Gao M, Wang W, Chen K, et al. Diabetic vascular diseases: molecular mechanisms and therapeutic strategies. Signal Transduction and Targeted Therapy. 2023;8(1):152.

59. Vinuesa A, Pomilio C, Gregosa A, Bentivegna M, Presa J, Bellotto M, et al. Inflammation and Insulin Resistance as Risk Factors and Potential Therapeutic Targets for Alzheimer’s Disease. Front Neurosci. 2021;15:653651.

60. Zhang Y, Chen H, Li R, Sterling K, Song W. Amyloid β-based therapy for Alzheimer’s disease: challenges, successes and future. Signal Transduction and Targeted Therapy. 2023;8(1):248.

