## Supplemental Materials for "Evaluating the causal association between type 2 diabetes and Alzheimer’s disease: a Two-Sample Mendelian Randomization Study"

**Supplementary Appendix 1. Search Strategy**

**Supplementary Appendix 2. Characters of the involved papers (among the EUR population)**

**Supplementary Appendix 3. Pooled results of meta-analysis for the included MR studies by using random effect model.**

**Supplementary Appendix 4. Information of instrumental variables consortiums**

**Supplementary Appendix 5. Information of instrumental variables of type 2 diabetes mellitus**

**Supplementary Appendix 8. Result of MR analysis between type 2 diabetes mellitus and Alzheimer’s disease (IGAP dataset)**

**Supplementary Appendix 9. Result of MR analysis between type 2 diabetes mellitus (T2DM) and Alzheimer’s disease (EDAB dataset)**

**Supplementary Appendix 10. Result of MR analysis between type 2 diabetes mellitus (T2DM) and Alzheimer’s disease (UKB dataset)**

**Supplementary Appendix 11. PRISMA checklist**

**Supplementary Appendix 12. STROBE-MR checklist of recommended items to address in reports of Mendelian randomization studies**

**Supplementary Appendix 1. Search Strategy**

Medline (PubMed), Web of Science, and Embase were searched to identify relevant studies. The keywords applied in the search among the three databases are similar. The search strategy for all databases is presented below.

| **Pubmed** | |
| --- | --- |
| #1 | "diabetes mellitus, type 2"[MeSH Terms] OR "type 2 diabetes"[Title/Abstract] OR "T2DM"[Title/Abstract] OR "T2D"[Title/Abstract] OR "NIDDM"[Title/Abstract] OR "non insulin dependent diabetes mellitus"[Title/Abstract] |
| #2 | "Alzheimer Disease"[MeSH Terms] OR "Alzheimer's disease"[Title/Abstract] OR "Alzheimer"[Title/Abstract] OR "Alzheimer's"[Title/Abstract] OR "AD"[Title/Abstract] OR "Dementia"[MeSH Terms] OR "Dementia"[Title/Abstract] |
| #3 | #1 AND #2 |
| #4 | "mendelian randomization analysis"[MeSH Terms] OR "mendelian randomisation"[Title/Abstract] OR "mendelian randomization"[Title/Abstract] OR "genetic instrumental variable"[Title/Abstract] OR "genetic instrument"[Title/Abstract] |
| #5 | (((Editorial[Publication Type]) OR (Letter[Publication Type])) OR (Case Reports[Publication Type])) OR (Comment[Publication Type]) OR (Congress[Publication Type]) OR (Review[Publication Type]) |
| #6 | (#3 AND #4) NOT #5 |
| **EMBASE** | |
| 1. 'diabetes mellitus, type 2'/exp OR 'type 2 diabetes':ti,ab OR 'T2DM':ti,ab OR 'T2D':ti,ab OR 'NIDDM':ti,ab OR 'non insulin dependent diabetes mellitus':ti,ab  2. 'alzheimer disease'/exp OR 'Alzheimer's disease':ti,ab OR 'Alzheimer':ti,ab OR 'Alzheimer's':ti,ab OR 'AD':ti,ab OR 'dementia'/exp OR 'dementia':ti,ab  3. 1 AND 2  4. 'mendelian randomization analysis'/exp OR 'mendelian randomisation':ti,ab OR 'mendelian randomization':ti,ab OR 'genetic instrumental variable':ti,ab OR 'genetic instrument':ti,ab  5. 3 AND 4  6. NOT ([editorial]/lim OR [letter]/lim OR [case report]/lim OR [conference abstract]/lim OR [review]/lim)  7. 5 AND 6 | |
| **Web of Science** | |
| TS=("diabetes mellitus, type 2" OR "type 2 diabetes" OR T2DM OR T2D OR NIDDM OR "non insulin dependent diabetes mellitus")  AND  TS=("Alzheimer Disease" OR "Alzheimer's disease" OR "Alzheimer" OR "Alzheimer's" OR AD OR "Dementia")  AND  TS=("mendelian randomization analysis" OR "mendelian randomisation" OR "mendelian randomization" OR "genetic instrumental variable" OR "genetic instrument")  NOT  DT=(Editorial OR Letter OR "Case Report" OR Comment OR Congress OR Review) | |

**Supplementary Appendix 2. Characters of the involved papers (among the EUR population)**

|  | **Exposure consortium** | **Outcome consortium** | **N** | **nSNP** | **MR** | **Result** |
| --- | --- | --- | --- | --- | --- | --- |
| Søren D. Østergaard et al, 2015 | Morris AP, Voight BF, Teslovich TM, et al. Large-scale association analysis provides insights into the genetic architecture and pathophysiology of type 2 diabetes. Nature genetics. 2012; 44(9):981. | Lambert JC, Ibrahim‐Verbaas CA, Harold D, Naj AC, Sims R, Bellenguez C, et al. Meta‐analysis of 74,046 individuals identifies 11 new susceptibility loci for Alzheimer’s disease. Nat Genet. 2013;45:1452–8. | 17,008/ 37,154; 8,572/11,312  (74,046) | 49 | 2SMR | OR [95% CI]: 1.01 [0.96–1.07]; p = 0.57 |
| Yuesong Pan et al, 2020 | Morris AP, Voight BF, Teslovich TM, et al. Large-scale association analysis provides insights into the genetic architecture and pathophysiology of type 2 diabetes. Nature genetics. 2012; 44(9):981.  Scott RA, Scott LJ, Mägi R, et al. An expanded genome-wide association study of type 2 diabetes in europeans. Diabetes. 2017; 66:2888–902. | Lambert JC, Ibrahim‐Verbaas CA, Harold D, Naj AC, Sims R, Bellenguez C, et al. Meta‐analysis of 74,046 individuals identifies 11 new susceptibility loci for Alzheimer’s disease. Nat Genet. 2013;45:1452–8. | 17,008/ 37,154; 8,572/11,312  (74,046) | 51 | 2SMR | OR [95% CI]: 1.02 (0.97-1.07), p = 0.52 |
| Jesper Qvist Thomassen et al, 2020 | Scott RA, Scott LJ, Mägi R, et al. An expanded genome-wide association study of type 2 diabetes in europeans. Diabetes. 2017; 66:2888–902. | Lambert JC, Ibrahim‐Verbaas CA, Harold D, Naj AC, Sims R, Bellenguez C, et al. Meta‐analysis of 74,046 individuals identifies 11 new susceptibility loci for Alzheimer’s disease. Nat Genet. 2013;45:1452–8. | 17,008/ 37,154; 8,572/11,312  (74,046) | 51 | 2SMR | OR [95% CI]: 1.04 (0.98–1.10) |
| Victoria Garfield et al, 2021 | Mahajan A, Taliun D, Thurner M, et al. Fine-mapping type 2 diabetes loci to single-variant resolution using high-density imputation and islet-specific epigenome maps. Nat Genet. 2018;50(11): 1505-1513. | Bycroft C, Freeman C, Petkova D, et al. The UK Biobank resource with deep phenotyping and genomic data. Nature 2018;562:203–209 | 488,377 | 157 | 2SMR | OR [95% CI]: 1.15 (0.87; 1.52) |
| Shea J. Andrews et al, 2021 | Xue A, Wu Y, Zhu Z, et al. Genome-wide association analyses identify 143 risk variants and putative regulatory mechanisms for type 2 diabetes. Nat Commun 2018;9:2941. | Kunkle BW, Grenier-Boley B, Sims R, et al. Genetic meta-analysis of diagnosed Alzheimer’s disease identifies new risk loci and implicates Aβ, tau, immunity and lipid processing. Nat Genet 2019;51:414–430. | 35,274/59,163  (94,437) | 218 | 2SMR | Beta (SE) 0.072 (0.017) |
| Lei Meng et al, 2022 | Mahajan A, Wessel J, Willems SM, Zhao W, Robertson NR, Chu AY, et al. Refining the accuracy of validated target identification through coding variant fine‐mapping in type 2 diabetes. Nat Genet. 2018;50:559–71. | Lambert JC, Ibrahim‐Verbaas CA, Harold D, Naj AC, Sims R, Bellenguez C, et al. Meta‐analysis of 74,046 individuals identifies 11 new susceptibility loci for Alzheimer’s disease. Nat Genet. 2013;45:1452–8. | 17,008/37,154; 8,572/11,312  (74,046) | 37 | 2SMR | OR [95% CI]: 1.34 (1.05, 1.70), p = 0.02 |
| Jiao Luo et al, 2023 | Vujkovic M, Keaton JM, Lynch JA, et al.; Discovery of 318 new risk loci for type 2 diabetes and related vascular outcomes among 1.4 million participants in a multi-ancestry meta-analysis. Nat Genet. 2020;52(7):680-691. | Bellenguez C, Küçükali F, Jansen IE, et al.; New insights into the genetic etiology of Alzheimer’s disease and related dementias. Nat Genet. 2022;54(4):412-436.  doi: 10.1038/s41588-022-01024-z | 111,326/677,663  (788,989) | 357 | 2SMR | OR [95% CI]: 1.02 (0.98-1.05) |
| Elin Dybjer et al, 2023 | Mahajan A, Taliun D, Thurner M, et al. Fine-mapping type 2 diabetes loci to single-variant resolution using high-density imputation and islet-specific epigenome maps. Nat Genet. 2018;50(11): 1505-1513. | Manjer J, Carlsson S, Elmståhl S, et al. The Malmö Diet and Cancer Study: Representativity, cancer incidence and mortality in participants and non-participants. Eur J Cancer Prev. 2001;10(6): 489-499. | 28,098/40, 807  (68,905) | 243 | 2SMR | Beta=-0.11, se=0.1, p = 0.259 |

2SMR, Two‐sample Mendelian randomization; 1SMR, One‐sample Mendelian randomization; IVW, Inverse variance weighting; T2D, type 2 diabetes; OR, odds ratio; CI, confidential interva

**Supplementary Appendix 3. Pooled results of meta-analysis for the included MR studies by using random effect model.**

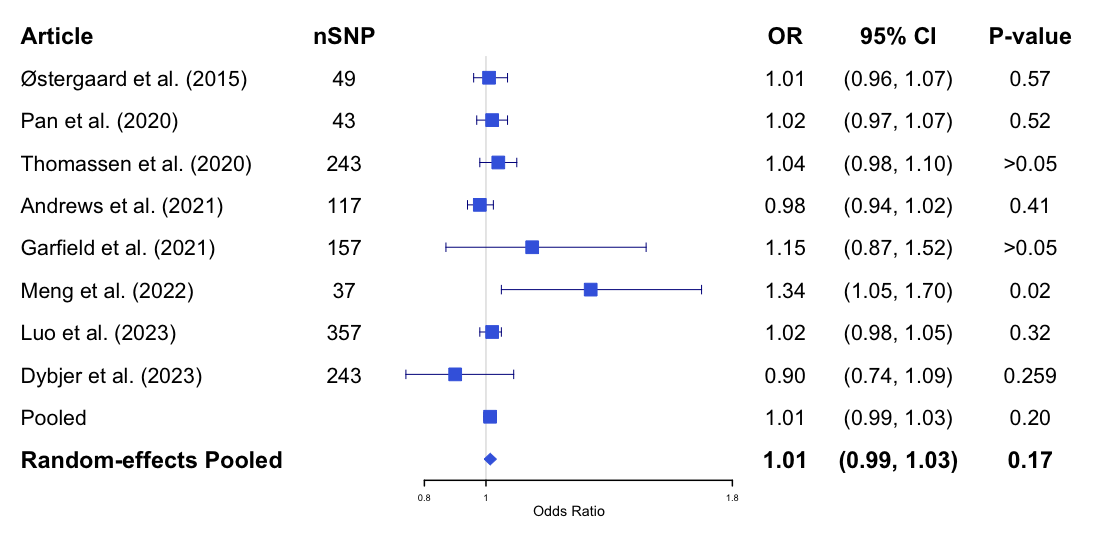

SNP, Single nucleotide polymorphism; OR, odds ratio; CI, confidence interval

**Supplementary Appendix 4. Information of instrumental variables consortiums**

DIAbetes Genetics Replication And Meta-analysis (DIAGRAM)

The DIAGRAM stage 1 meta-analyses comprises 26,676 T2D case and 132,532 control subjects from 18 studies genotyped using commercial genome-wide single nucleotide variant (SNV) arrays. The Metabochip stage 2 follow-up comprises 14,545 T2D case and 38,994 control subjects (*N*_eff_ = 38,645) from 16 nonoverlapping stage 1 studies (D2D2007, DANISH, DIAGEN, DILGOM, DRsEXTRA, EMIL-Ulm, FUSION2, NHR, IMPROVE, InterACT-CMC, Leipzig, METSIM, HUNT/TROMSO, SCARFSHEEP, STR, Warren2/58BC). Additional follow-up was conducted in 2,796 T2D case and 4,601 control subjects from the European Prospective Investigation into Cancer and Nutrition-InterAct (EPIC-InterAct) study and in 9,747 T2D case and 61,857 control subjects from the Resource for Genetic Epidemiology on Adult Health and Aging (GERA) study (1). 13 novel T2D-associated loci (*P* < 5 × 10^-8^) were identified, including variants near the *GLP2R*, *GIP*, and *HLA-DQA1* genes (1). Various definitions of type 2 diabetes mellitus used across included studies, commonly based on diagnostic criteria such as fasting glucose (≥ 7.0 mmol/L), HbA1c (≥ 6.5%), or non-fasting glucose (≥ 11.1 mmol/L). Diagnostic methods include self-reported physician diagnoses, medical records, hospital discharge data, and electronic health registries. Some definitions also require treatment with diabetes medication. Specific criteria like the exclusion of cases with markers for type 1 diabetes (e.g., antibodies) or the use of American Diabetes Association and WHO standards are also mentioned. Controls were typically individuals with glucose levels below diagnostic thresholds and no diabetes diagnosis or treatment history, often matched to cases by demographics and health metrics (1).

DIAbetes Meta-ANalysis of Trans-Ethnic association studies (DIAMANTE)

The DIAMANTE Consortium was established to assemble T2DM GWAS across diverse ancestry groups. Analyses of the European and East Asian ancestry components of DIAMANTE have previously been reported. In the GWAS included in current study, coding variant data was aggregated for 81,412 type 2 diabetes cases and 370,832 controls of diverse ancestry through performing both European-specific (EUR) and trans-ethnic (TE) meta-analyses, identifying 40 coding variant association signals (P < 2.2 × 10-7); of these, 16 map outside known risk-associated loci.  Genotypes were assembled from: (a) 58,425 cases and 188,032 controls genotyped with the exome-array; (b) 14,608 cases and 174,322 controls from UK Biobank and GERA (Genetic Epidemiology on Adult Health and Aging) genotyped with GWAS arrays enriched for exome content and/or coverage of low-frequency variation across ethnic groups; and (c) 8,379 cases and 8,478 controls with whole-exome sequence from GoT2D/T2D-GENES and SIGMA studies (2-6).  T2D diagnosis criteria across studies generally include prior T2D diagnosis, fasting glucose levels equal to or exceeding 7 mmol/L, HbA1c levels above 6.5%, or the use of glucose-lowering medications. For prevalent and incident cases, some studies use criteria from established guidelines such as the American Diabetes Association, WHO, or cohort-specific frameworks (e.g., CHS, CMS). In some settings, the criteria extend to 2-hour glucose tests (values above 11.1 mmol/L) or casual glucose measurements when fasting data is unavailable. Diagnosis sources vary from self-reported cases, physician assessments, medication records, or laboratory-confirmed hyperglycemia, with additional specifications in some cohorts, such as C-peptide levels or the absence of anti-GAD antibodies (2).

Another latest multi-ancestry meta-analysis was also applied in the included MR studies. Type 2 diabetes genetic susceptibility via multi-ancestry meta-analysis of 228,499 cases and 1,178,783 controls in the Million Veteran Program (MVP) and other studies with non-overlapping participants: DIAMANTE Consortium, Penn Medicine Biobank, Pakistan Genomic Resource, Biobank Japan, Malmö Diet and Cancer Study, Medstar, and PennCath was investigated. In this study, 568 associations, including 286 autosomal, 7 X-chromosomal and 25 identified in ancestry-specific analyses that were previously unreported was reported. The diagnosis of type 2 diabets varies from clinical and administrative criteria, including physician diagnosis, self-reports, to biochemical measures. Core diagnostic criteria involve fasting glucose levels ≥ 7.0 mmol/L, HbA1c ≥ 6.5%, and/or the use of glucose-lowering medications. Some studies rely on specific diagnostic codes (such as ICD-9) or criteria from the WHO or ADA. Advanced protocols include exclusion of Type 1 diabetes through age criteria, antibody testing (e.g., GAD antibodies), and C-peptide levels, while others apply a multi-source approach, verifying diagnosis across electronic health records, registries, or hospital records (7).

**Supplementary Appendix 5. Information of instrumental variables of type 2 diabetes mellitus**

| SNP | chr | pos | A1 | A2 | eaf | beta | se | pval | gene |
| --- | --- | --- | --- | --- | --- | --- | --- | --- | --- |
| rs12741141 | 1 | 6669970 | G | C | 0.362 | 0.040 | 0.005 | 1.37E-16 | KLHL21 |
| rs4845987 | 1 | 11306279 | C | G | 0.692 | 0.028 | 0.005 | 2.78E-08 | MTOR |
| rs10916784 | 1 | 20729451 | G | C | 0.585 | 0.027 | 0.005 | 3.91E-09 | LINC01141 |
| rs6685701 | 1 | 26868639 | A | G | 0.279 | 0.030 | 0.005 | 1.14E-08 | RPS6KA1 |
| rs111824905 | 1 | 28110797 | C | T | 0.055 | 0.069 | 0.012 | 4.02E-09 | STX12 |
| rs10915188 | 1 | 29024956 | A | G | 0.585 | 0.029 | 0.005 | 3.69E-10 | GMEB1 |
| rs61779275 | 1 | 39820310 | T | C | 0.220 | 0.075 | 0.006 | 3.35E-43 | MACF1 |
| rs3176466 | 1 | 51438365 | C | T | 0.903 | 0.064 | 0.008 | 9.36E-16 | CDKN2C |
| rs2269247 | 1 | 64107284 | C | T | 0.814 | 0.035 | 0.006 | 5.74E-09 | PGM1 |
| rs10889560 | 1 | 65989878 | A | C | 0.088 | 0.047 | 0.008 | 4.01E-09 | LEPR |
| rs4655617 | 1 | 67010654 | C | A | 0.434 | 0.028 | 0.005 | 2.45E-09 | SGIP1 |
| rs2613503 | 1 | 72839774 | A | C | 0.801 | 0.039 | 0.006 | 7.48E-11 | RNU6-1246P,RPL31P12 |
| rs197379 | 1 | 112292303 | C | T | 0.387 | 0.027 | 0.005 | 9.14E-09 | INKA2 |
| rs1127215 | 1 | 117532790 | C | T | 0.585 | 0.043 | 0.005 | 1.93E-20 | PTGFRN |
| rs41276588 | 1 | 118148384 | A | G | 0.283 | 0.038 | 0.005 | 4.10E-13 | TENT5C-DT |
| rs1493694 | 1 | 120526982 | T | C | 0.107 | 0.071 | 0.007 | 6.51E-22 | NOTCH2 |
| rs72692805 | 1 | 149894355 | G | A | 0.924 | 0.054 | 0.009 | 6.01E-10 | SV2A,SF3B4 |
| rs145904381 | 1 | 151017991 | T | C | 0.990 | 0.174 | 0.023 | 4.31E-14 | BNIPL |
| rs1194606 | 1 | 154294260 | C | T | 0.233 | 0.030 | 0.006 | 3.41E-08 | AQP10 |
| rs3020781 | 1 | 155269776 | G | A | 0.270 | 0.033 | 0.005 | 1.05E-10 | PKLR |
| rs4916253 | 1 | 172361032 | G | T | 0.431 | 0.027 | 0.005 | 3.09E-09 | PIGC,DNM3 |
| rs539515 | 1 | 177889025 | C | A | 0.200 | 0.051 | 0.008 | 1.20E-10 | NA |
| rs567185 | 1 | 201763499 | T | C | 0.634 | 0.037 | 0.005 | 1.55E-14 | NAV1,IPO9-AS1 |
| rs12048743 | 1 | 205114873 | G | C | 0.440 | 0.032 | 0.005 | 2.11E-12 | DSTYK |
| rs7538321 | 1 | 205789455 | T | A | 0.125 | 0.042 | 0.007 | 1.33E-09 | PM20D1-AS1 |
| rs2336938 | 1 | 206618799 | A | C | 0.484 | 0.030 | 0.005 | 9.54E-11 | SRGAP2 |
| rs79687284 | 1 | 214150821 | C | G | 0.035 | 0.148 | 0.020 | 2.60E-16 | PROX1 |
| rs3738430 | 1 | 214157546 | G | A | 0.976 | 0.130 | 0.021 | 2.80E-09 | NA |
| rs340874 | 1 | 214159256 | C | T | 0.553 | 0.067 | 0.005 | 5.37E-48 | PROX1,PROX1-AS1 |
| rs2820444 | 1 | 219741820 | G | A | 0.713 | 0.047 | 0.005 | 8.66E-21 | ZC3H11B |
| rs348330 | 1 | 229672955 | G | A | 0.369 | 0.053 | 0.007 | 6.50E-16 | ABCB10 |
| rs291367 | 1 | 235690800 | G | A | 0.630 | 0.044 | 0.007 | 6.10E-10 | NA |
| rs62107261 | 2 | 422144 | T | C | 0.954 | 0.113 | 0.016 | 3.80E-12 | TMEM18 |
| rs10188334 | 2 | 653874 | C | T | 0.828 | 0.050 | 0.006 | 7.76E-16 | TMEM18,LINC01875 |
| rs11680058 | 2 | 16574669 | A | G | 0.838 | 0.056 | 0.010 | 1.23E-08 | CYRIA,GACAT3 |
| rs7558413 | 2 | 18721662 | A | G | 0.570 | 0.029 | 0.005 | 6.44E-10 | KCNS3,RDH14 |
| rs34845373 | 2 | 25635771 | A | G | 0.729 | 0.037 | 0.005 | 1.18E-12 | DTNB |
| rs72803684 | 2 | 26192802 | T | C | 0.046 | 0.069 | 0.012 | 6.01E-09 | KIF3C |
| rs1260326 | 2 | 27730940 | C | T | 0.595 | 0.064 | 0.005 | 2.55E-42 | GCKR |
| rs4952673 | 2 | 43423870 | A | G | 0.470 | 0.039 | 0.006 | 1.60E-09 | NA |
| rs13414140 | 2 | 43671176 | C | T | 0.886 | 0.118 | 0.007 | 4.30E-60 | THADA |
| rs10193538 | 2 | 58981064 | T | G | 0.610 | 0.039 | 0.007 | 8.90E-09 | BNIPL |
| rs980183 | 2 | 59311536 | G | A | 0.394 | 0.036 | 0.005 | 1.02E-14 | LINC01122,LINC01793 |
| rs243018 | 2 | 60586707 | G | C | 0.463 | 0.056 | 0.005 | 1.52E-33 | MIR4432HG |
| rs2540949 | 2 | 65284231 | A | T | 0.616 | 0.050 | 0.005 | 9.19E-26 | CEP68 |
| rs2028150 | 2 | 65655012 | C | G | 0.598 | 0.049 | 0.007 | 2.30E-12 | CEP68 |
| rs1430780 | 2 | 67878328 | T | C | 0.323 | 0.027 | 0.005 | 3.35E-08 | LINC02831,LINC01812 |
| rs34506349 | 2 | 100598726 | G | A | 0.960 | 0.068 | 0.012 | 1.76E-08 | AFF3 |
| rs17624303 | 2 | 105148418 | C | T | 0.725 | 0.029 | 0.005 | 4.88E-08 | LINC01102 |
| rs72836348 | 2 | 111888043 | G | A | 0.892 | 0.056 | 0.008 | 3.20E-13 | BCL2L11,MIR4435-2HG |
| rs34589210 | 2 | 112795492 | A | G | 0.143 | 0.039 | 0.007 | 6.89E-09 | TMEM87B,MERTK |
| rs9784137 | 2 | 121325908 | G | A | 0.849 | 0.061 | 0.007 | 5.25E-21 | Y_RNA,LINC01101 |
| rs2033159 | 2 | 145261174 | C | A | 0.234 | 0.035 | 0.006 | 4.90E-10 | ZEB2 |
| rs7609422 | 2 | 146348037 | G | A | 0.410 | 0.030 | 0.005 | 2.67E-10 | RPL6P5,METAP2P1 |
| rs7559658 | 2 | 147920213 | C | T | 0.190 | 0.034 | 0.006 | 3.17E-09 | LINC01911,RNU6-692P |
| rs13020443 | 2 | 152167830 | C | T | 0.498 | 0.031 | 0.005 | 2.65E-11 | NMI,TNFAIP6 |
| rs7568172 | 2 | 158335340 | G | A | 0.940 | 0.067 | 0.010 | 2.64E-12 | CYTIP |
| rs6432613 | 2 | 161145612 | G | A | 0.724 | 0.039 | 0.005 | 6.73E-14 | RBMS1 |
| rs13389219 | 2 | 165528876 | C | T | 0.603 | 0.065 | 0.005 | 7.35E-44 | COBLL1 |
| rs12992995 | 2 | 175197545 | C | A | 0.732 | 0.031 | 0.005 | 2.46E-09 | SP9,LINC01305 |
| rs6715901 | 2 | 179650954 | G | A | 0.512 | 0.027 | 0.005 | 3.06E-09 | TTN |
| rs6741676 | 2 | 181618654 | A | G | 0.668 | 0.032 | 0.005 | 3.64E-11 | SCHLAP1 |
| rs12463719 | 2 | 203450680 | A | G | 0.282 | 0.032 | 0.005 | 6.15E-10 | BMPR2,MTCO1P17 |
| rs34329895 | 2 | 208870017 | A | G | 0.397 | 0.028 | 0.005 | 3.10E-09 | PLEKHM3 |
| rs13005841 | 2 | 212302573 | A | T | 0.713 | 0.029 | 0.005 | 1.89E-08 | ERBB4 |
| rs17354348 | 2 | 213835977 | A | G | 0.742 | 0.029 | 0.005 | 2.99E-08 | MIR4776-1,IKZF2 |
| rs6736415 | 2 | 226851035 | C | T | 0.180 | 0.047 | 0.008 | 9.30E-09 | NA |
| rs2972145 | 2 | 227101309 | C | T | 0.635 | 0.090 | 0.005 | 7.45E-80 | MIR5702,NYAP2 |
| rs77402945 | 2 | 227225798 | C | T | 0.850 | 0.051 | 0.009 | 9.00E-09 | NA |
| rs7561798 | 2 | 228973660 | G | A | 0.485 | 0.028 | 0.005 | 1.16E-09 | SPHKAP |
| rs838735 | 2 | 234324192 | C | G | 0.384 | 0.029 | 0.005 | 3.64E-10 | DGKD |
| rs3872707 | 3 | 9514016 | A | G | 0.140 | 0.045 | 0.007 | 3.46E-11 | SETD5 |
| rs9826367 | 3 | 12294202 | A | G | 0.560 | 0.041 | 0.006 | 2.90E-10 | NA |
| rs17036160 | 3 | 12329783 | C | T | 0.880 | 0.103 | 0.007 | 3.04E-47 | PPARG |
| rs4465929 | 3 | 15741389 | T | C | 0.404 | 0.030 | 0.005 | 5.11E-11 | ANKRD28,BTD |
| rs35352848 | 3 | 23455582 | T | C | 0.793 | 0.062 | 0.006 | 7.37E-28 | UBE2E2 |
| rs118109589 | 3 | 23635623 | G | A | 0.030 | 0.120 | 0.018 | 7.40E-11 | NA |
| rs10490871 | 3 | 35667761 | G | A | 0.369 | 0.027 | 0.005 | 6.58E-09 | ARPP21,RNU6-243P |
| rs11129735 | 3 | 36870230 | A | G | 0.462 | 0.026 | 0.005 | 1.53E-08 | TRANK1 |
| rs11926707 | 3 | 46925539 | C | T | 0.626 | 0.046 | 0.008 | 1.69E-08 | PTH1R |
| rs62262091 | 3 | 47693664 | T | C | 0.089 | 0.056 | 0.009 | 1.14E-10 | SMARCC1 |
| rs4688760 | 3 | 49980596 | T | C | 0.674 | 0.034 | 0.005 | 1.10E-11 | RBM6 |
| rs2581787 | 3 | 53127677 | T | G | 0.558 | 0.025 | 0.005 | 3.14E-08 | RFT1 |
| rs76263492 | 3 | 54828827 | T | G | 0.045 | 0.091 | 0.016 | 6.30E-09 | NA |
| rs2292662 | 3 | 63897215 | C | T | 0.844 | 0.056 | 0.007 | 7.64E-18 | SCAANT1,ATXN7 |
| rs4132228 | 3 | 64708114 | C | T | 0.703 | 0.047 | 0.005 | 6.22E-21 | ADAMTS9-AS2 |
| rs844215 | 3 | 71656045 | C | T | 0.587 | 0.026 | 0.005 | 2.04E-08 | FOXP1-DT,EIF4E3 |
| rs11922794 | 3 | 72813582 | C | G | 0.251 | 0.030 | 0.005 | 1.46E-08 | SHQ1 |
| rs13085136 | 3 | 72865183 | C | T | 0.928 | 0.077 | 0.012 | 1.50E-08 | SHQ1 |
| rs1437055 | 3 | 86831077 | A | C | 0.613 | 0.027 | 0.005 | 1.19E-08 | VGLL3,LINC02070 |
| rs11716527 | 3 | 89986280 | C | T | 0.095 | 0.049 | 0.009 | 6.69E-09 | MTCO1P6,U3 |
| rs6438247 | 3 | 115084080 | C | T | 0.155 | 0.044 | 0.007 | 4.05E-11 | GAP43,EIF4E2P2 |
| rs11708067 | 3 | 123065778 | A | G | 0.771 | 0.078 | 0.006 | 1.50E-46 | ADCY5 |
| rs9873519 | 3 | 124921457 | T | C | 0.534 | 0.038 | 0.005 | 3.57E-16 | SLC12A8 |
| rs9828772 | 3 | 129333182 | C | G | 0.899 | 0.052 | 0.008 | 2.70E-11 | PLXND1,CARMIL2P1 |
| rs1225052 | 3 | 131644937 | G | A | 0.371 | 0.027 | 0.005 | 1.00E-08 | CPNE4 |
| rs9852406 | 3 | 135625498 | T | C | 0.254 | 0.038 | 0.005 | 8.96E-13 | SDHBP1,EPHB1 |
| rs6766859 | 3 | 138055136 | C | T | 0.376 | 0.033 | 0.005 | 7.23E-12 | MRAS,NME9 |
| rs73872717 | 3 | 141134569 | C | T | 0.953 | 0.086 | 0.011 | 4.78E-15 | ZBTB38 |
| rs34573045 | 3 | 149196752 | G | C | 0.420 | 0.031 | 0.005 | 2.27E-11 | TM4SF4 |
| rs62271373 | 3 | 150066540 | A | T | 0.056 | 0.069 | 0.010 | 4.44E-11 | TSC22D2 |
| rs7619041 | 3 | 152095371 | T | A | 0.487 | 0.043 | 0.008 | 2.76E-08 | NA |
| rs74672008 | 3 | 152451616 | G | A | 0.960 | 0.080 | 0.012 | 1.38E-11 | ATP5MGP5,MBNL1 |
| rs56394279 | 3 | 160171092 | C | T | 0.468 | 0.031 | 0.005 | 5.30E-12 | TRIM59,B3GAT3P1 |
| rs1449348 | 3 | 168225055 | C | T | 0.859 | 0.045 | 0.007 | 6.64E-12 | EGFEM1P |
| rs9873618 | 3 | 170733076 | G | A | 0.709 | 0.058 | 0.005 | 2.41E-30 | SLC2A2 |
| rs686998 | 3 | 173119768 | G | A | 0.539 | 0.027 | 0.005 | 6.11E-09 | NLGN1 |
| rs2313211 | 3 | 183738626 | T | A | 0.448 | 0.029 | 0.005 | 4.06E-10 | ABCC5,EEF1A1P8 |
| rs10937208 | 3 | 184877626 | G | A | 0.136 | 0.044 | 0.007 | 1.06E-10 | EHHADH-AS1,C3orf70 |
| rs9854769 | 3 | 185520948 | G | A | 0.319 | 0.108 | 0.005 | 1.06E-107 | IGF2BP2 |
| rs113672528 | 3 | 185540817 | C | A | 0.020 | 0.140 | 0.021 | 1.10E-10 | NA |
| rs3887925 | 3 | 186665645 | T | C | 0.547 | 0.068 | 0.007 | 3.10E-22 | ST6GAL1 |
| rs6777684 | 3 | 187741842 | G | A | 0.608 | 0.057 | 0.005 | 3.77E-33 | LINC01991,LPP-AS2 |
| rs7619708 | 3 | 195810187 | T | C | 0.758 | 0.033 | 0.005 | 3.46E-10 | TFRC,LINC00885 |
| rs1531583 | 4 | 744972 | T | G | 0.045 | 0.099 | 0.011 | 7.06E-19 | PCGF3 |
| rs72501964 | 4 | 1267203 | G | T | 0.961 | 0.083 | 0.013 | 2.09E-10 | CTBP1-DT |
| rs56337234 | 4 | 1784403 | C | T | 0.505 | 0.041 | 0.005 | 1.88E-18 | FGFR3,TACC3 |
| rs362307 | 4 | 3241845 | T | C | 0.075 | 0.050 | 0.009 | 3.77E-08 | HTT |
| rs10937721 | 4 | 6306763 | C | G | 0.588 | 0.084 | 0.005 | 6.31E-70 | WFS1,PPP2R2C |
| rs2011603 | 4 | 18025484 | A | G | 0.725 | 0.038 | 0.005 | 1.59E-13 | LCORL |
| rs11940813 | 4 | 20210953 | G | A | 0.137 | 0.037 | 0.007 | 2.65E-08 | SLIT2,RPL21P46 |
| rs10938398 | 4 | 45186139 | A | G | 0.431 | 0.043 | 0.005 | 3.91E-20 | PRDX4P1,THAP12P9 |
| rs62310934 | 4 | 48880627 | C | G | 0.611 | 0.030 | 0.005 | 1.37E-10 | OCIAD1,OCIAD2 |
| rs17086692 | 4 | 53134293 | G | T | 0.687 | 0.047 | 0.008 | 2.48E-08 | SPATA18 |
| rs114447556 | 4 | 53207093 | T | C | 0.083 | 0.058 | 0.009 | 7.79E-11 | RNU6-1252P,SPATA18 |
| rs2055997 | 4 | 76535086 | G | A | 0.701 | 0.031 | 0.005 | 6.53E-10 | CDKL2 |
| rs11723275 | 4 | 77528821 | C | A | 0.467 | 0.027 | 0.005 | 4.26E-09 | SHROOM3 |
| rs10471048 | 4 | 83587562 | G | C | 0.349 | 0.034 | 0.005 | 2.31E-12 | SCD5 |
| rs7660000 | 4 | 89751858 | C | T | 0.713 | 0.031 | 0.005 | 1.29E-09 | FAM13A |
| rs7656001 | 4 | 91243865 | A | G | 0.550 | 0.026 | 0.005 | 1.23E-08 | CCSER1 |
| rs6821438 | 4 | 95091911 | A | G | 0.531 | 0.029 | 0.005 | 3.66E-10 | SMARCAD1-DT |
| rs3755879 | 4 | 96114385 | A | G | 0.315 | 0.033 | 0.005 | 5.78E-11 | UNC5C |
| rs7695096 | 4 | 103932556 | C | T | 0.521 | 0.038 | 0.005 | 1.27E-16 | SLC9B1 |
| rs17035289 | 4 | 106048291 | C | T | 0.171 | 0.043 | 0.006 | 5.70E-12 | TET2,RNU6-351P |
| rs11098676 | 4 | 123833154 | C | T | 0.788 | 0.054 | 0.010 | 2.03E-08 | NUDT6 |
| rs12509379 | 4 | 129179458 | T | G | 0.212 | 0.031 | 0.006 | 2.66E-08 | PGRMC2,LARP1B |
| rs1724557 | 4 | 137094048 | C | A | 0.418 | 0.025 | 0.005 | 4.73E-08 | RNU1-89P,TERF1P3 |
| rs12505942 | 4 | 140906390 | T | C | 0.662 | 0.030 | 0.005 | 7.03E-10 | MAML3 |
| rs75686861 | 4 | 145621328 | A | G | 0.093 | 0.047 | 0.008 | 4.16E-09 | AC098588.2,HHIP |
| rs6819331 | 4 | 153504295 | C | T | 0.680 | 0.040 | 0.005 | 2.21E-16 | RPS3AP18,TMEM154 |
| rs28819812 | 4 | 157652753 | C | A | 0.675 | 0.038 | 0.005 | 3.36E-14 | LINC02272,PDGFC |
| rs72695645 | 4 | 185713608 | G | A | 0.860 | 0.061 | 0.007 | 8.66E-20 | ACSL1 |
| rs35901985 | 4 | 186580062 | A | G | 0.824 | 0.035 | 0.006 | 1.69E-08 | SORBS2 |
| rs17250977 | 5 | 14753745 | G | A | 0.038 | 0.113 | 0.016 | 2.00E-11 | ANKH |
| rs6885132 | 5 | 14768092 | C | G | 0.900 | 0.078 | 0.011 | 9.50E-13 | NA |
| rs1061813 | 5 | 14847331 | G | A | 0.463 | 0.043 | 0.007 | 3.37E-09 | ANKH |
| rs114136102 | 5 | 36084426 | C | T | 0.040 | 0.072 | 0.012 | 2.05E-09 | LMBRD2,UGT3A2 |
| rs13155752 | 5 | 44680687 | C | A | 0.397 | 0.032 | 0.005 | 1.12E-11 | LINC02224,RN7SL383P |
| rs152839 | 5 | 50145266 | C | T | 0.576 | 0.026 | 0.005 | 1.25E-08 | PARP8,LINC02106 |
| rs12187734 | 5 | 51763665 | C | T | 0.523 | 0.029 | 0.005 | 2.24E-10 | MFSD4BP1,RPS17P11 |
| rs3811978 | 5 | 52100489 | G | A | 0.170 | 0.053 | 0.009 | 4.20E-10 | NA |
| rs4865796 | 5 | 53272664 | A | G | 0.682 | 0.047 | 0.005 | 4.64E-21 | ARL15 |
| rs256904 | 5 | 55810305 | T | A | 0.730 | 0.069 | 0.005 | 2.55E-39 | C5orf67 |
| rs9687832 | 5 | 55861595 | A | G | 0.198 | 0.077 | 0.009 | 1.70E-20 | ANKRD55 |
| rs4976033 | 5 | 67714246 | G | A | 0.411 | 0.028 | 0.005 | 1.93E-09 | PIK3R1 |
| rs253412 | 5 | 74955841 | A | G | 0.652 | 0.046 | 0.005 | 2.77E-21 | ANKDD1B |
| rs6878122 | 5 | 76427311 | G | A | 0.311 | 0.055 | 0.005 | 4.60E-29 | PDE8B,ZBED3-AS1 |
| rs12519500 | 5 | 78436905 | C | A | 0.654 | 0.038 | 0.005 | 1.76E-15 | DMGDH |
| rs7719891 | 5 | 86577352 | G | A | 0.261 | 0.040 | 0.007 | 8.96E-09 | RASA1 |
| rs2410767 | 5 | 87705268 | C | G | 0.779 | 0.033 | 0.006 | 6.79E-09 | LINC02060,TMEM161B-DT |
| rs145510090 | 5 | 101273694 | A | T | 0.053 | 0.100 | 0.015 | 1.10E-12 | NA |
| rs62369303 | 5 | 101711308 | C | T | 0.280 | 0.047 | 0.007 | 3.70E-11 | NA |
| rs17154859 | 5 | 102290912 | T | G | 0.320 | 0.044 | 0.007 | 1.40E-10 | NA |
| rs75432112 | 5 | 102586407 | A | G | 0.048 | 0.134 | 0.011 | 5.34E-36 | PPIP5K2,MACIR |
| rs10077431 | 5 | 112927686 | C | A | 0.785 | 0.049 | 0.009 | 4.76E-08 | YTHDC2 |
| rs329122 | 5 | 133864599 | A | G | 0.427 | 0.026 | 0.005 | 1.25E-08 | JADE2 |
| rs112667817 | 5 | 137823156 | C | T | 0.879 | 0.061 | 0.011 | 5.76E-09 | RPL7P19,ETF1 |
| rs890940 | 5 | 158026744 | T | C | 0.211 | 0.048 | 0.006 | 9.78E-18 | EBF1,LINC02227 |
| rs9379084 | 6 | 7231843 | G | A | 0.880 | 0.075 | 0.008 | 3.70E-22 | RREB1 |
| rs9505086 | 6 | 7232186 | C | T | 0.380 | 0.048 | 0.007 | 2.70E-13 | NA |
| rs727734 | 6 | 15475051 | A | T | 0.748 | 0.030 | 0.005 | 3.06E-08 | JARID2 |
| rs9368112 | 6 | 19718157 | T | C | 0.525 | 0.026 | 0.005 | 2.69E-08 | LNC-LBCS |
| rs11964747 | 6 | 20485898 | C | T | 0.810 | 0.060 | 0.008 | 5.10E-13 | NA |
| rs7756992 | 6 | 20679709 | G | A | 0.271 | 0.122 | 0.005 | 5.52E-128 | CDKAL1 |
| rs72832338 | 6 | 20775412 | A | G | 0.081 | 0.090 | 0.012 | 9.90E-15 | NA |
| rs4077404 | 6 | 20876613 | G | A | 0.780 | 0.061 | 0.008 | 5.30E-15 | NA |
| rs3094682 | 6 | 31264461 | C | A | 0.806 | 0.060 | 0.006 | 2.37E-23 | HLA-C |
| rs2246618 | 6 | 31478986 | T | C | 0.307 | 0.051 | 0.008 | 1.20E-09 | NA |
| rs2844492 | 6 | 31518169 | G | A | 0.030 | 0.140 | 0.022 | 1.10E-10 | NA |
| rs1063355 | 6 | 32627714 | G | T | 0.602 | 0.071 | 0.008 | 3.72E-19 | HLA-DQB1 |
| rs9275184 | 6 | 32654714 | C | T | 0.111 | 0.098 | 0.010 | 5.59E-24 | MTCO3P1,HLA-DQB1 |
| rs9296095 | 6 | 33542523 | T | C | 0.803 | 0.034 | 0.006 | 4.75E-09 | GGNBP1,BAK1 |
| rs10305420 | 6 | 39016636 | C | T | 0.616 | 0.032 | 0.005 | 2.69E-11 | GLP1R |
| rs34298980 | 6 | 40409243 | T | C | 0.501 | 0.038 | 0.007 | 4.20E-09 | LRFN2 |
| rs4714422 | 6 | 41012405 | G | A | 0.239 | 0.029 | 0.005 | 4.80E-08 | OARD1 |
| rs11967262 | 6 | 43760327 | G | C | 0.488 | 0.037 | 0.005 | 2.23E-15 | VEGFA,LINC02537 |
| rs10456526 | 6 | 43814625 | A | G | 0.290 | 0.051 | 0.007 | 5.00E-13 | NA |
| rs3798519 | 6 | 50788778 | C | A | 0.180 | 0.050 | 0.006 | 5.49E-17 | TFAP2B |
| rs1819564 | 6 | 51505337 | A | T | 0.027 | 0.076 | 0.014 | 3.85E-08 | PKHD1 |
| rs60519666 | 6 | 107427166 | G | A | 0.677 | 0.035 | 0.005 | 3.19E-12 | BEND3 |
| rs55812705 | 6 | 111738793 | T | C | 0.749 | 0.031 | 0.005 | 7.45E-09 | MFSD4B,REV3L |
| rs72951506 | 6 | 118011723 | C | T | 0.852 | 0.042 | 0.007 | 9.22E-11 | NUS1 |
| rs2008027 | 6 | 126052359 | G | A | 0.504 | 0.027 | 0.005 | 4.28E-09 | HEY2-AS1 |
| rs11759026 | 6 | 126792095 | G | A | 0.231 | 0.065 | 0.007 | 6.68E-19 | CENPW,MIR588 |
| rs12194820 | 6 | 127401978 | A | T | 0.765 | 0.046 | 0.006 | 9.19E-17 | RPS4XP9,RSPO3 |
| rs7739842 | 6 | 131954797 | G | T | 0.194 | 0.033 | 0.006 | 1.98E-08 | ENPP3 |
| rs1573090 | 6 | 137302159 | T | G | 0.535 | 0.045 | 0.005 | 3.87E-22 | NHEG1,RPL35AP3 |
| rs11155073 | 6 | 139837128 | T | C | 0.421 | 0.030 | 0.005 | 4.43E-11 | ATP5PBP6,LINC01625 |
| rs197482 | 6 | 143069315 | C | T | 0.616 | 0.030 | 0.005 | 2.64E-10 | HIVEP2,ADGRG6 |
| rs9383649 | 6 | 153428102 | G | A | 0.419 | 0.033 | 0.005 | 1.06E-12 | RGS17 |
| rs543159 | 6 | 160776017 | C | A | 0.520 | 0.032 | 0.005 | 1.29E-12 | SLC22A3 |
| rs4709746 | 6 | 164133001 | C | T | 0.869 | 0.058 | 0.007 | 1.03E-16 | QKI |
| rs4721089 | 7 | 1872921 | T | C | 0.757 | 0.034 | 0.006 | 7.92E-10 | MAD1L1 |
| rs798549 | 7 | 2760750 | C | A | 0.282 | 0.030 | 0.005 | 1.46E-08 | AMZ1 |
| rs62450857 | 7 | 4683258 | A | G | 0.133 | 0.039 | 0.007 | 1.85E-08 | FOXK1,CYP3A54P |
| rs13237518 | 7 | 12269593 | A | C | 0.414 | 0.029 | 0.005 | 7.72E-10 | TMEM106B |
| rs1122518 | 7 | 13900325 | C | T | 0.473 | 0.026 | 0.005 | 1.25E-08 | RBMX2P4,ETV1 |
| rs17168486 | 7 | 14898282 | T | C | 0.181 | 0.068 | 0.007 | 2.30E-17 | DGKB |
| rs2191349 | 7 | 15064309 | T | G | 0.539 | 0.066 | 0.005 | 6.41E-48 | AGMO,GTF3AP5 |
| rs38221 | 7 | 15926228 | T | C | 0.260 | 0.033 | 0.005 | 6.62E-10 | CRPPA,RPL36AP26 |
| rs583769 | 7 | 18331915 | A | G | 0.251 | 0.031 | 0.005 | 2.69E-09 | HDAC9 |
| rs75693095 | 7 | 23440057 | C | G | 0.021 | 0.112 | 0.017 | 1.42E-11 | IGF2BP3 |
| rs62451127 | 7 | 27971300 | G | A | 0.945 | 0.086 | 0.014 | 2.20E-09 | NA |
| rs1513272 | 7 | 28200097 | C | T | 0.509 | 0.081 | 0.005 | 3.97E-71 | JAZF1 |
| rs917195 | 7 | 30728452 | C | T | 0.769 | 0.047 | 0.006 | 2.71E-17 | CRHR2 |
| rs17439448 | 7 | 40816653 | T | C | 0.121 | 0.040 | 0.007 | 2.98E-08 | SUGCT |
| rs2268576 | 7 | 44189023 | C | T | 0.490 | 0.039 | 0.006 | 1.40E-09 | NA |
| rs2908286 | 7 | 44234737 | T | C | 0.171 | 0.068 | 0.006 | 2.64E-29 | GCK |
| rs12539264 | 7 | 48839003 | G | A | 0.282 | 0.029 | 0.005 | 6.62E-09 | GDI2P1,LINC02838 |
| rs2876826 | 7 | 50581972 | G | A | 0.215 | 0.031 | 0.006 | 3.33E-08 | DDC |
| rs2103132 | 7 | 69782073 | C | G | 0.251 | 0.032 | 0.005 | 1.88E-09 | AUTS2 |
| rs67755137 | 7 | 74108135 | A | G | 0.190 | 0.033 | 0.006 | 2.24E-08 | GTF2I-AS1,GTF2I |
| rs10240790 | 7 | 89880949 | G | A | 0.704 | 0.028 | 0.005 | 4.69E-08 | CFAP69 |
| rs534043 | 7 | 100312724 | G | A | 0.874 | 0.045 | 0.007 | 8.03E-10 | POP7,EPO |
| rs1968204 | 7 | 102800137 | T | C | 0.107 | 0.057 | 0.008 | 1.08E-12 | RPL23AP95,DPY19L2P2 |
| rs39328 | 7 | 103444978 | T | C | 0.427 | 0.028 | 0.005 | 3.29E-09 | RELN |
| rs13239186 | 7 | 117510621 | T | C | 0.302 | 0.054 | 0.009 | 2.70E-10 | CTTNBP2 |
| rs1562398 | 7 | 130457931 | G | C | 0.417 | 0.041 | 0.005 | 1.99E-18 | KLF14,LINC-PINT |
| rs62492368 | 7 | 150537635 | A | G | 0.315 | 0.034 | 0.005 | 6.38E-12 | AOC1 |
| rs6459733 | 7 | 156930550 | G | C | 0.666 | 0.051 | 0.005 | 9.66E-25 | UBE3C,MNX1-AS1 |
| rs117173251 | 8 | 4186731 | T | C | 0.033 | 0.077 | 0.014 | 2.48E-08 | CSMD1 |
| rs6984305 | 8 | 9178268 | A | T | 0.110 | 0.061 | 0.010 | 2.10E-09 | NA |
| rs34990153 | 8 | 9996389 | A | G | 0.561 | 0.038 | 0.005 | 1.99E-16 | MSRA |
| rs2409742 | 8 | 11069960 | C | T | 0.507 | 0.036 | 0.005 | 7.34E-15 | LINC00529 |
| rs12056338 | 8 | 12643055 | T | G | 0.414 | 0.031 | 0.005 | 2.88E-11 | LINC03019 |
| rs17294565 | 8 | 14124809 | C | A | 0.381 | 0.027 | 0.005 | 4.74E-09 | SGCZ |
| rs10096633 | 8 | 19830921 | C | T | 0.880 | 0.070 | 0.010 | 8.70E-13 | NA |
| rs1059592 | 8 | 22477778 | A | G | 0.354 | 0.027 | 0.005 | 2.40E-08 | CCAR2 |
| rs17818197 | 8 | 25872634 | G | A | 0.212 | 0.035 | 0.006 | 4.99E-10 | EBF2 |
| rs2725371 | 8 | 30854033 | A | G | 0.311 | 0.037 | 0.005 | 2.14E-13 | PURG |
| rs1060731 | 8 | 41435225 | T | C | 0.290 | 0.042 | 0.007 | 3.50E-09 | NA |
| rs13262861 | 8 | 41508577 | C | A | 0.824 | 0.102 | 0.006 | 2.84E-59 | NKX6-3,ANK1 |
| rs148766658 | 8 | 41552046 | C | T | 0.040 | 0.110 | 0.017 | 7.90E-11 | NA |
| rs2241896 | 8 | 41555473 | T | C | 0.630 | 0.046 | 0.007 | 4.70E-11 | NA |
| rs62515938 | 8 | 57483013 | T | C | 0.261 | 0.029 | 0.005 | 2.97E-08 | LINC00968,RPL37P6 |
| rs11786992 | 8 | 95685147 | A | C | 0.640 | 0.040 | 0.007 | 1.70E-09 | NA |
| rs10097617 | 8 | 95961626 | T | C | 0.480 | 0.037 | 0.005 | 1.93E-16 | NDUFAF6 |
| rs34340810 | 8 | 105661926 | G | C | 0.927 | 0.054 | 0.009 | 4.08E-10 | ZFPM2 |
| rs4734193 | 8 | 110140564 | C | A | 0.532 | 0.034 | 0.006 | 3.70E-08 | NUDCD1,TRHR |
| rs2737226 | 8 | 116639474 | T | C | 0.389 | 0.038 | 0.005 | 2.52E-16 | TRPS1 |
| rs11558471 | 8 | 118185733 | A | G | 0.684 | 0.103 | 0.005 | 3.98E-98 | SLC30A8 |
| rs17772814 | 8 | 128711742 | G | A | 0.915 | 0.075 | 0.013 | 3.44E-09 | CASC11 |
| rs1561927 | 8 | 129568078 | C | T | 0.282 | 0.035 | 0.005 | 8.61E-12 | LINC00824 |
| rs3757969 | 8 | 145551199 | G | C | 0.376 | 0.048 | 0.005 | 3.60E-22 | DGAT1,SCRT1 |
| rs2294120 | 8 | 146003567 | A | G | 0.544 | 0.044 | 0.008 | 1.62E-08 | ZNF34 |
| rs756145 | 9 | 1039939 | A | G | 0.309 | 0.029 | 0.005 | 5.52E-09 | H3P29,LINC01230 |
| rs10974438 | 9 | 4291928 | C | A | 0.356 | 0.047 | 0.005 | 8.98E-23 | GLIS3 |
| rs10963942 | 9 | 19080352 | G | A | 0.397 | 0.037 | 0.005 | 4.49E-15 | HAUS6 |
| rs7867635 | 9 | 20241069 | C | T | 0.411 | 0.037 | 0.006 | 4.21E-09 | MLLT3,SLC24A2 |
| rs1063192 | 9 | 22003367 | A | G | 0.560 | 0.058 | 0.006 | 2.90E-19 | NA |
| rs10811660 | 9 | 22134068 | G | A | 0.828 | 0.239 | 0.010 | 1.40E-115 | CDKN2A/B |
| rs7018475 | 9 | 22137685 | G | T | 0.270 | 0.110 | 0.007 | 6.30E-48 | NA |
| rs11793831 | 9 | 23362311 | T | G | 0.405 | 0.027 | 0.005 | 4.21E-09 | LINC01239,SUMO2P2 |
| rs1412234 | 9 | 28410683 | C | T | 0.319 | 0.044 | 0.005 | 4.00E-19 | LINGO2 |
| rs12001437 | 9 | 34074476 | C | T | 0.373 | 0.034 | 0.005 | 6.27E-13 | DCAF12,RN7SKP114 |
| rs1929883 | 9 | 81344701 | G | A | 0.580 | 0.039 | 0.006 | 4.84E-10 | PSAT1,MTND2P8 |
| rs17791513 | 9 | 81905590 | A | G | 0.930 | 0.100 | 0.013 | 2.90E-14 | NA |
| rs2796441 | 9 | 84308948 | G | A | 0.591 | 0.059 | 0.005 | 9.96E-37 | TLE1-DT |
| rs555784 | 9 | 85318704 | T | A | 0.616 | 0.030 | 0.005 | 1.70E-10 | MTCO3P40,RPS6P12 |
| rs10821311 | 9 | 96943059 | A | G | 0.319 | 0.036 | 0.005 | 4.00E-13 | MIRLET7A1HG,LINC02603 |
| rs7046845 | 9 | 97804641 | A | C | 0.910 | 0.047 | 0.008 | 2.26E-08 | AOPEP |
| rs7858727 | 9 | 111936128 | C | A | 0.223 | 0.034 | 0.006 | 8.33E-10 | EPB41L4B |
| rs1431819 | 9 | 116943357 | G | A | 0.689 | 0.029 | 0.005 | 6.33E-09 | COL27A1 |
| rs7026688 | 9 | 125975397 | G | A | 0.863 | 0.044 | 0.007 | 3.40E-11 | STRBP |
| rs1752169 | 9 | 126586563 | A | C | 0.270 | 0.032 | 0.005 | 1.64E-09 | DENND1A |
| rs495203 | 9 | 136145240 | T | C | 0.335 | 0.049 | 0.005 | 1.03E-24 | ABO |
| rs448918 | 9 | 136885979 | A | G | 0.287 | 0.033 | 0.005 | 1.11E-09 | BRD3OS,VAV2 |
| rs28429551 | 9 | 139243334 | A | T | 0.740 | 0.073 | 0.006 | 9.51E-40 | GPSM1 |
| rs11257655 | 10 | 12307894 | T | C | 0.219 | 0.091 | 0.007 | 7.25E-35 | CDC123,RN7SL198P |
| rs878017 | 10 | 13566204 | A | G | 0.541 | 0.035 | 0.006 | 1.13E-08 | BEND7 |
| rs36051838 | 10 | 34018730 | C | T | 0.089 | 0.044 | 0.008 | 4.54E-08 | LINC02628,LINC00838 |
| rs12263348 | 10 | 65305252 | T | C | 0.339 | 0.028 | 0.005 | 7.77E-09 | REEP3 |
| rs10998304 | 10 | 70342775 | C | T | 0.452 | 0.031 | 0.005 | 1.77E-11 | TET1 |
| rs177045 | 10 | 71321279 | G | A | 0.316 | 0.068 | 0.007 | 6.60E-18 | NEUROG3 |
| rs2642588 | 10 | 71466578 | G | T | 0.702 | 0.049 | 0.007 | 2.20E-14 | NEUROG3 |
| rs827237 | 10 | 72648336 | T | C | 0.205 | 0.037 | 0.006 | 3.56E-10 | SGPL1 |
| rs2675662 | 10 | 75599127 | A | G | 0.564 | 0.027 | 0.005 | 8.30E-09 | CAMK2G |
| rs7099048 | 10 | 77647107 | A | G | 0.504 | 0.028 | 0.005 | 8.93E-10 | LRMDA |
| rs703981 | 10 | 80942855 | G | C | 0.544 | 0.061 | 0.005 | 2.13E-40 | ZMIZ1 |
| rs11201999 | 10 | 88124501 | C | T | 0.537 | 0.026 | 0.005 | 1.25E-08 | GRID1 |
| rs10788575 | 10 | 89768584 | A | G | 0.149 | 0.035 | 0.006 | 3.71E-08 | PTEN,MED6P1 |
| rs7071943 | 10 | 93956552 | G | T | 0.653 | 0.044 | 0.005 | 2.22E-19 | CPEB3 |
| rs7084673 | 10 | 94167087 | A | G | 0.390 | 0.046 | 0.007 | 3.20E-12 | NA |
| rs77014180 | 10 | 94307157 | G | A | 0.060 | 0.089 | 0.014 | 9.60E-11 | NA |
| rs1111875 | 10 | 94462882 | C | T | 0.592 | 0.092 | 0.005 | 5.21E-89 | HHEX,Y_RNA |
| rs146935743 | 10 | 94466064 | C | T | 0.970 | 0.170 | 0.030 | 4.00E-08 | HHEX/IDE |
| rs10882891 | 10 | 99059645 | C | A | 0.415 | 0.031 | 0.005 | 1.68E-11 | Metazoa_SRP,RPL12P27 |
| rs2862954 | 10 | 101912064 | T | C | 0.517 | 0.029 | 0.005 | 1.56E-10 | ERLIN1 |
| rs2250301 | 10 | 104548393 | G | A | 0.748 | 0.032 | 0.005 | 1.27E-09 | WBP1L |
| rs10787287 | 10 | 112647195 | T | C | 0.759 | 0.036 | 0.006 | 6.56E-11 | PDCD4 |
| rs1927157 | 10 | 114635381 | C | T | 0.760 | 0.064 | 0.008 | 2.90E-17 | NA |
| rs114222749 | 10 | 114668249 | T | C | 0.023 | 0.170 | 0.021 | 9.20E-16 | NA |
| rs116425039 | 10 | 114681965 | G | A | 0.990 | 0.280 | 0.036 | 1.00E-14 | NA |
| rs116859590 | 10 | 114752410 | T | C | 0.026 | 0.240 | 0.021 | 3.70E-29 | NA |
| rs34872471 | 10 | 114754071 | C | T | 0.300 | 0.310 | 0.007 | 5.63E-144 | NA |
| rs78025551 | 10 | 114757956 | C | G | 0.850 | 0.150 | 0.009 | 8.40E-63 | NA |
| rs61872774 | 10 | 114765390 | A | G | 0.013 | 0.280 | 0.030 | 2.80E-21 | NA |
| rs10885404 | 10 | 114773068 | G | T | 0.820 | 0.150 | 0.018 | 6.50E-17 | TCF7L2 |
| rs141241414 | 10 | 114775551 | A | G | 0.990 | 0.190 | 0.028 | 3.50E-11 | NA |
| rs116369954 | 10 | 114793572 | C | T | 0.030 | 0.290 | 0.018 | 2.10E-56 | NA |
| rs11196201 | 10 | 114803307 | T | A | 0.080 | 0.180 | 0.012 | 9.70E-54 | NA |
| rs4918791 | 10 | 114830306 | G | A | 0.630 | 0.074 | 0.007 | 2.20E-25 | NA |
| rs6585206 | 10 | 114859251 | A | G | 0.190 | 0.066 | 0.008 | 9.40E-16 | NA |
| rs11196229 | 10 | 114866172 | G | A | 0.750 | 0.092 | 0.008 | 6.90E-34 | NA |
| rs10885419 | 10 | 114893956 | G | C | 0.270 | 0.057 | 0.007 | 2.40E-15 | NA |
| rs1225404 | 10 | 114914665 | T | C | 0.640 | 0.060 | 0.007 | 5.00E-19 | NA |
| rs2280141 | 10 | 124193181 | T | G | 0.520 | 0.045 | 0.006 | 1.09E-13 | PLEKHA1,ARMS2 |
| rs4929965 | 11 | 2197286 | A | G | 0.385 | 0.062 | 0.005 | 2.46E-38 | ASCL2,MIR4686 |
| rs231361 | 11 | 2691500 | A | G | 0.256 | 0.077 | 0.007 | 5.00E-25 | KCNQ1 |
| rs2283220 | 11 | 2755548 | A | G | 0.690 | 0.049 | 0.007 | 1.40E-09 | KCNQ1 |
| rs2237895 | 11 | 2857194 | C | A | 0.423 | 0.073 | 0.005 | 2.73E-54 | KCNQ1 |
| rs2237897 | 11 | 2858546 | C | T | 0.954 | 0.207 | 0.017 | 8.40E-32 | KCNQ1 |
| rs10769936 | 11 | 8654528 | C | T | 0.707 | 0.035 | 0.005 | 6.29E-12 | TRIM66 |
| rs2403221 | 11 | 9852475 | A | G | 0.657 | 0.032 | 0.005 | 5.60E-11 | SBF2 |
| rs117316450 | 11 | 14518419 | G | C | 0.019 | 0.131 | 0.018 | 2.07E-13 | COPB1 |
| rs5219 | 11 | 17409572 | T | C | 0.373 | 0.069 | 0.005 | 3.15E-48 | KCNJ11 |
| rs62618693 | 11 | 32956492 | C | T | 0.957 | 0.085 | 0.011 | 6.83E-14 | QSER1 |
| rs11555762 | 11 | 43876698 | T | C | 0.305 | 0.041 | 0.005 | 8.65E-17 | HSD17B12 |
| rs11038672 | 11 | 45846498 | C | G | 0.476 | 0.029 | 0.005 | 1.04E-10 | SLC35C1,CRY2 |
| rs7124681 | 11 | 47529947 | A | C | 0.410 | 0.037 | 0.006 | 6.40E-09 | NA |
| rs7929543 | 11 | 49351026 | C | A | 0.083 | 0.083 | 0.014 | 2.20E-09 | TYRL |
| rs116861182 | 11 | 55588216 | C | A | 0.055 | 0.064 | 0.011 | 5.90E-09 | OR5D18,OR5L2 |
| rs174541 | 11 | 61565908 | T | C | 0.646 | 0.029 | 0.005 | 8.63E-10 | FADS2 |
| rs35169799 | 11 | 64031241 | T | C | 0.066 | 0.050 | 0.009 | 4.79E-08 | PLCB3 |
| rs1783541 | 11 | 65294799 | T | C | 0.203 | 0.048 | 0.006 | 3.89E-17 | SCYL1 |
| rs144245804 | 11 | 69453044 | G | A | 0.973 | 0.130 | 0.015 | 1.13E-17 | CCND1,LINC01488 |
| rs11602873 | 11 | 72460762 | A | T | 0.844 | 0.098 | 0.006 | 8.91E-52 | ARAP1 |
| rs480840 | 11 | 74625997 | C | T | 0.423 | 0.025 | 0.005 | 4.46E-08 | XRRA1 |
| rs10899283 | 11 | 76505202 | C | T | 0.771 | 0.031 | 0.006 | 1.46E-08 | TSKU |
| rs10830963 | 11 | 92708710 | G | C | 0.278 | 0.089 | 0.005 | 4.57E-68 | MTNR1B |
| rs57235767 | 11 | 93013531 | C | T | 0.710 | 0.046 | 0.007 | 6.60E-11 | NA |
| rs10893829 | 11 | 128042575 | T | C | 0.853 | 0.058 | 0.010 | 1.30E-10 | ETS1 |
| rs10750397 | 11 | 128234144 | A | G | 0.290 | 0.048 | 0.005 | 1.01E-20 | LINC02098,ETS1 |
| rs11221333 | 11 | 128383687 | T | C | 0.220 | 0.053 | 0.008 | 4.20E-12 | NA |
| rs11063029 | 12 | 4301301 | T | C | 0.057 | 0.089 | 0.014 | 1.70E-10 | NA |
| rs11063069 | 12 | 4374373 | G | A | 0.210 | 0.056 | 0.008 | 7.40E-13 | NA |
| rs3217792 | 12 | 4384696 | C | T | 0.913 | 0.113 | 0.011 | 2.60E-21 | CCND2 |
| rs76895963 | 12 | 4384844 | T | G | 0.980 | 0.482 | 0.027 | 1.40E-69 | CCND2 |
| rs3217860 | 12 | 4399050 | G | A | 0.258 | 0.049 | 0.007 | 3.90E-09 | CCND2 |
| rs12299509 | 12 | 4406281 | G | A | 0.479 | 0.047 | 0.007 | 2.09E-10 | CCND2 |
| rs67013744 | 12 | 6681786 | G | A | 0.160 | 0.035 | 0.006 | 4.11E-08 | CHD4 |
| rs10841868 | 12 | 21781246 | G | T | 0.739 | 0.032 | 0.005 | 1.65E-09 | GYS2,LDHB |
| rs11048458 | 12 | 26465585 | T | C | 0.250 | 0.046 | 0.005 | 4.66E-18 | ITPR2-AS1 |
| rs10771372 | 12 | 27962260 | C | T | 0.805 | 0.072 | 0.006 | 2.20E-35 | RN7SKP15,PTHLH |
| rs10771813 | 12 | 31367856 | C | A | 0.544 | 0.026 | 0.005 | 2.60E-08 | DDX11,RPL13AP22 |
| rs10844518 | 12 | 33410780 | G | A | 0.287 | 0.033 | 0.005 | 8.80E-11 | ASS1P14,SYT10 |
| rs2733289 | 12 | 41838235 | C | T | 0.478 | 0.030 | 0.005 | 3.98E-11 | PDZRN4 |
| rs11181613 | 12 | 43046449 | C | A | 0.855 | 0.043 | 0.007 | 1.37E-10 | LINC02451 |
| rs2732480 | 12 | 48736303 | C | A | 0.565 | 0.034 | 0.005 | 3.05E-13 | ZNF641 |
| rs7132908 | 12 | 50263148 | A | G | 0.390 | 0.033 | 0.005 | 1.41E-12 | FAIM2 |
| rs1872635 | 12 | 54541750 | A | G | 0.686 | 0.028 | 0.005 | 1.28E-08 | SMUG1 |
| rs2583921 | 12 | 66170481 | C | A | 0.092 | 0.095 | 0.008 | 8.98E-33 | RPSAP52 |
| rs1042725 | 12 | 66358347 | T | C | 0.490 | 0.054 | 0.006 | 1.60E-17 | NA |
| rs1705263 | 12 | 71523043 | C | A | 0.569 | 0.040 | 0.005 | 1.22E-17 | TSPAN8 |
| rs11107116 | 12 | 93978504 | T | G | 0.220 | 0.047 | 0.009 | 3.75E-08 | SOCS2 |
| rs11108094 | 12 | 95928113 | A | C | 0.068 | 0.060 | 0.009 | 1.16E-10 | USP44 |
| rs2197973 | 12 | 95928560 | T | C | 0.538 | 0.039 | 0.007 | 3.60E-08 | USP44 |
| rs113036477 | 12 | 97848227 | C | T | 0.938 | 0.072 | 0.010 | 4.09E-13 | RMST |
| rs3764002 | 12 | 108618630 | C | T | 0.741 | 0.040 | 0.005 | 2.69E-14 | WSCD2 |
| rs34965774 | 12 | 118412373 | A | G | 0.138 | 0.052 | 0.007 | 1.38E-14 | RFC5,KSR2 |
| rs117389214 | 12 | 121119057 | C | T | 0.050 | 0.088 | 0.015 | 6.50E-09 | NA |
| rs73226260 | 12 | 121380541 | G | A | 0.967 | 0.120 | 0.019 | 7.00E-11 | NA |
| rs1800574 | 12 | 121416864 | T | C | 0.030 | 0.160 | 0.019 | 8.90E-17 | NA |
| rs56348580 | 12 | 121432117 | G | C | 0.696 | 0.058 | 0.005 | 2.33E-30 | HNF1A |
| rs12820906 | 12 | 123493123 | A | G | 0.756 | 0.043 | 0.006 | 2.21E-15 | PITPNM2 |
| rs12823740 | 12 | 124458002 | C | A | 0.661 | 0.041 | 0.005 | 1.48E-17 | ZNF664,RFLNA |
| rs825476 | 12 | 124568456 | T | C | 0.581 | 0.052 | 0.007 | 6.80E-13 | ZNF664-FAM101A |
| rs11830243 | 12 | 132544694 | T | C | 0.113 | 0.044 | 0.007 | 2.10E-09 | EP400 |
| rs11614914 | 12 | 133070294 | T | C | 0.325 | 0.039 | 0.005 | 4.65E-15 | FBRSL1 |
| rs12305809 | 12 | 133777466 | G | A | 0.607 | 0.033 | 0.005 | 1.45E-12 | ZNF268 |
| rs314879 | 13 | 23309382 | C | T | 0.226 | 0.039 | 0.006 | 5.74E-12 | DDX39AP1,SNORD36 |
| rs34584161 | 13 | 26776999 | A | G | 0.761 | 0.052 | 0.005 | 3.54E-22 | RNF6 |
| rs9319382 | 13 | 28245127 | C | T | 0.679 | 0.028 | 0.005 | 2.87E-08 | POLR1D |
| rs3742305 | 13 | 31036642 | C | G | 0.729 | 0.030 | 0.005 | 1.51E-08 | HMGB1 |
| rs576674 | 13 | 33554302 | G | A | 0.185 | 0.061 | 0.006 | 1.25E-23 | KL,TOMM22P3 |
| rs4397977 | 13 | 41688401 | A | G | 0.344 | 0.029 | 0.005 | 3.05E-09 | RN7SL597P,MIR3168 |
| rs9316500 | 13 | 51094114 | T | G | 0.708 | 0.047 | 0.005 | 9.97E-21 | DLEU7,DLEU1 |
| rs9563574 | 13 | 58656599 | T | C | 0.823 | 0.041 | 0.006 | 1.64E-11 | RNA5SP30,LINC02338 |
| rs9563615 | 13 | 59077406 | A | T | 0.710 | 0.049 | 0.007 | 6.40E-11 | SRGAP2D |
| rs11616380 | 13 | 80705315 | G | T | 0.721 | 0.079 | 0.005 | 7.99E-54 | LINC01080,SPRY2 |
| rs1475655 | 13 | 91963080 | A | T | 0.747 | 0.044 | 0.005 | 2.47E-16 | PPIAP23,MIR17HG |
| rs9555581 | 13 | 109944192 | C | T | 0.610 | 0.030 | 0.005 | 3.72E-10 | LINC00370 |
| rs8005994 | 14 | 29744532 | A | G | 0.655 | 0.027 | 0.005 | 4.52E-08 | RNU11-5P,LINC02326 |
| rs17522122 | 14 | 33302882 | T | G | 0.476 | 0.034 | 0.005 | 1.27E-13 | AKAP6 |
| rs799661 | 14 | 35390146 | C | T | 0.871 | 0.043 | 0.007 | 7.11E-09 | IGBP1P1,BAZ1A-AS1 |
| rs8018512 | 14 | 38818723 | G | A | 0.742 | 0.037 | 0.005 | 1.30E-12 | KRT8P1,CLEC14A |
| rs2933211 | 14 | 47313541 | A | G | 0.501 | 0.027 | 0.005 | 6.24E-09 | MDGA2 |
| rs10137475 | 14 | 58797953 | G | A | 0.426 | 0.026 | 0.005 | 4.43E-08 | ARID4A |
| rs4899280 | 14 | 69526307 | T | C | 0.330 | 0.028 | 0.005 | 1.02E-08 | DCAF5 |
| rs8008540 | 14 | 74948180 | C | T | 0.569 | 0.030 | 0.005 | 7.99E-11 | NPC2 |
| rs2056857 | 14 | 77300863 | C | T | 0.594 | 0.026 | 0.005 | 2.59E-08 | LRRC74A |
| rs10145154 | 14 | 79939525 | T | C | 0.216 | 0.055 | 0.006 | 1.06E-22 | NRXN3 |
| rs8010382 | 14 | 91963722 | G | A | 0.437 | 0.032 | 0.005 | 5.74E-12 | PPP4R3A |
| rs73347525 | 14 | 101255172 | A | G | 0.810 | 0.049 | 0.008 | 7.36E-09 | MEG3 |
| rs12890750 | 14 | 103860309 | G | T | 0.639 | 0.028 | 0.005 | 2.84E-09 | MARK3 |
| rs11073147 | 15 | 36392562 | G | A | 0.540 | 0.025 | 0.005 | 4.26E-08 | LINC02853,COX6CP4 |
| rs8032939 | 15 | 38834033 | C | T | 0.250 | 0.043 | 0.007 | 8.40E-09 | NA |
| rs34715063 | 15 | 38873115 | C | T | 0.124 | 0.095 | 0.012 | 2.30E-19 | RASGRP1 |
| rs11639470 | 15 | 39639171 | C | G | 0.547 | 0.027 | 0.005 | 1.34E-08 | LINC02915,THBS1 |
| rs484943 | 15 | 40398754 | T | C | 0.318 | 0.033 | 0.005 | 7.61E-11 | BMF |
| rs2289739 | 15 | 41801512 | T | G | 0.350 | 0.050 | 0.007 | 1.57E-14 | LTK |
| rs74804697 | 15 | 52588722 | C | G | 0.957 | 0.084 | 0.012 | 4.68E-12 | MYO5C,MYO5A |
| rs75332279 | 15 | 53099306 | C | T | 0.096 | 0.056 | 0.008 | 4.93E-12 | RPSAP55,ONECUT1 |
| rs2435907 | 15 | 57333416 | A | G | 0.589 | 0.029 | 0.005 | 1.32E-09 | TCF12 |
| rs8033609 | 15 | 60938816 | A | C | 0.545 | 0.027 | 0.005 | 1.11E-08 | RORA |
| rs7163757 | 15 | 62391608 | C | T | 0.574 | 0.041 | 0.005 | 1.64E-18 | NPM1P47,C2CD4B |
| rs34143602 | 15 | 63940058 | G | A | 0.422 | 0.035 | 0.005 | 1.00E-13 | HERC1 |
| rs1874832 | 15 | 67260238 | G | A | 0.168 | 0.038 | 0.007 | 3.22E-09 | SMASR,SMAD3-DT |
| rs4776970 | 15 | 68080886 | A | T | 0.638 | 0.029 | 0.005 | 6.14E-10 | MAP2K5 |
| rs12917449 | 15 | 74331659 | C | A | 0.197 | 0.036 | 0.006 | 4.71E-10 | PML |
| rs6495182 | 15 | 75814388 | C | T | 0.748 | 0.041 | 0.005 | 2.90E-14 | PTPN9 |
| rs12910361 | 15 | 77782335 | G | A | 0.698 | 0.072 | 0.005 | 2.97E-44 | LINGO1,HMG20A |
| rs36111056 | 15 | 83461873 | G | A | 0.780 | 0.034 | 0.006 | 1.97E-09 | FSD2 |
| rs8031576 | 15 | 90380214 | C | A | 0.289 | 0.057 | 0.005 | 1.74E-29 | ARPIN-AP3S2,AP3S2 |
| rs2290203 | 15 | 91512067 | A | G | 0.202 | 0.056 | 0.006 | 1.79E-22 | PRC1,PRC1-AS1 |
| rs55857387 | 16 | 300388 | T | C | 0.805 | 0.052 | 0.006 | 6.71E-19 | FAM234A |
| rs4984980 | 16 | 968292 | A | G | 0.188 | 0.035 | 0.006 | 3.79E-09 | LMF1 |
| rs12933120 | 16 | 3634746 | A | C | 0.139 | 0.042 | 0.007 | 3.18E-10 | SLX4 |
| rs9927842 | 16 | 15153717 | T | C | 0.162 | 0.038 | 0.007 | 6.84E-09 | PDXDC1 |
| rs62034975 | 16 | 20392415 | C | G | 0.300 | 0.031 | 0.005 | 6.60E-10 | PDILT |
| rs7188071 | 16 | 28917644 | T | C | 0.364 | 0.029 | 0.005 | 1.33E-09 | RABEP2 |
| rs8054556 | 16 | 29958216 | A | G | 0.462 | 0.036 | 0.005 | 3.68E-15 | TMEM219 |
| rs7203521 | 16 | 53769293 | A | G | 0.610 | 0.041 | 0.007 | 3.30E-10 | NA |
| rs1421085 | 16 | 53800954 | C | T | 0.414 | 0.118 | 0.005 | 5.63E-144 | FTO |
| rs62033401 | 16 | 53814470 | C | T | 0.870 | 0.057 | 0.010 | 5.00E-09 | NA |
| rs6499646 | 16 | 53843533 | T | C | 0.920 | 0.070 | 0.012 | 4.80E-09 | NA |
| rs2032912 | 16 | 69568303 | G | T | 0.584 | 0.042 | 0.005 | 2.11E-19 | CYB5B,NFAT5 |
| rs72802342 | 16 | 75234872 | C | A | 0.921 | 0.115 | 0.009 | 5.58E-38 | ZFP1,CTRB2 |
| rs2925979 | 16 | 81534790 | T | C | 0.308 | 0.045 | 0.005 | 1.54E-19 | CMIP |
| rs11646052 | 16 | 85716463 | G | A | 0.392 | 0.026 | 0.005 | 2.61E-08 | GINS2 |
| rs11117364 | 16 | 88132199 | G | A | 0.666 | 0.029 | 0.005 | 7.71E-09 | LINC02182,BANP |
| rs12920022 | 16 | 89564055 | A | T | 0.160 | 0.039 | 0.007 | 2.26E-09 | SPG7 |
| rs11870735 | 17 | 481604 | T | C | 0.182 | 0.034 | 0.006 | 1.68E-08 | VPS53 |
| rs8071043 | 17 | 3988451 | C | T | 0.328 | 0.054 | 0.005 | 5.16E-28 | ZZEF1 |
| rs2243102 | 17 | 4839149 | C | T | 0.422 | 0.026 | 0.005 | 4.94E-08 | SLC25A11,GP1BA |
| rs858519 | 17 | 7531965 | T | C | 0.444 | 0.026 | 0.005 | 1.88E-08 | SHBG |
| rs7219033 | 17 | 9787958 | A | G | 0.314 | 0.029 | 0.005 | 4.94E-09 | GLP2R |
| rs2297508 | 17 | 17715317 | C | G | 0.368 | 0.033 | 0.005 | 1.86E-12 | SREBF1 |
| rs7220340 | 17 | 27566326 | A | G | 0.455 | 0.027 | 0.005 | 4.00E-09 | TWF1P1,CRYBA1 |
| rs12602834 | 17 | 29637308 | G | A | 0.391 | 0.029 | 0.005 | 7.09E-10 | EVI2B,NF1 |
| rs4796224 | 17 | 34842521 | G | A | 0.474 | 0.025 | 0.005 | 2.71E-08 | ZNHIT3 |
| rs11657964 | 17 | 36100767 | A | G | 0.404 | 0.059 | 0.005 | 1.23E-36 | HNF1B |
| rs11078916 | 17 | 37746307 | T | C | 0.296 | 0.037 | 0.005 | 4.84E-13 | NEUROD2,CDK12 |
| rs684214 | 17 | 40696915 | T | C | 0.273 | 0.042 | 0.005 | 2.57E-16 | HSD17B1P1,NAGLU |
| rs9900074 | 17 | 46124326 | G | C | 0.925 | 0.055 | 0.009 | 3.24E-10 | NFE2L1-DT |
| rs35895680 | 17 | 47060322 | C | A | 0.686 | 0.056 | 0.005 | 1.42E-28 | GIP |
| rs1451506 | 17 | 57407019 | A | G | 0.127 | 0.042 | 0.008 | 2.78E-08 | YPEL2,SNRPGP17 |
| rs4325 | 17 | 61563200 | C | A | 0.539 | 0.035 | 0.005 | 6.92E-14 | ACE |
| rs17631783 | 17 | 61687600 | C | T | 0.737 | 0.049 | 0.009 | 3.95E-08 | TACO1 |
| rs11655898 | 17 | 62201374 | C | T | 0.070 | 0.059 | 0.010 | 5.85E-10 | ERN1 |
| rs2080090 | 17 | 65828371 | A | T | 0.188 | 0.053 | 0.006 | 7.84E-19 | BPTF |
| rs61736066 | 17 | 70645032 | G | A | 0.917 | 0.051 | 0.009 | 2.08E-09 | SLC39A11 |
| rs1656794 | 17 | 75386909 | G | A | 0.716 | 0.031 | 0.005 | 3.58E-09 | SEPTIN9 |
| rs62075585 | 17 | 76762039 | G | A | 0.474 | 0.030 | 0.005 | 1.25E-10 | CYTH1 |
| rs9912236 | 17 | 77895311 | C | T | 0.750 | 0.031 | 0.006 | 1.56E-08 | LINC01979,LINC01978 |
| rs7240767 | 18 | 7070642 | C | T | 0.379 | 0.037 | 0.006 | 5.03E-09 | LAMA1 |
| rs11662800 | 18 | 13271367 | A | G | 0.418 | 0.028 | 0.005 | 2.42E-09 | LDLRAD4 |
| rs303760 | 18 | 21083738 | T | C | 0.348 | 0.034 | 0.005 | 1.48E-12 | RMC1 |
| rs346240 | 18 | 40063830 | G | A | 0.223 | 0.031 | 0.006 | 3.62E-08 | LINC00907 |
| rs72926932 | 18 | 53050646 | C | A | 0.078 | 0.075 | 0.008 | 7.58E-20 | TCF4 |
| rs17684074 | 18 | 54675384 | G | C | 0.739 | 0.031 | 0.005 | 4.12E-09 | WDR7 |
| rs1517037 | 18 | 56878274 | C | T | 0.813 | 0.038 | 0.006 | 3.22E-10 | SEC11C,GRP |
| rs663640 | 18 | 57846077 | T | C | 0.218 | 0.050 | 0.006 | 1.15E-19 | RNU4-17P,MC4R |
| rs12454712 | 18 | 60845884 | T | C | 0.617 | 0.041 | 0.005 | 8.30E-18 | BCL2 |
| rs2658746 | 18 | 74582340 | C | T | 0.390 | 0.030 | 0.005 | 9.89E-11 | ZNF236 |
| rs35004890 | 19 | 1224286 | T | G | 0.221 | 0.036 | 0.006 | 8.57E-10 | STK11 |
| rs12977104 | 19 | 4949921 | A | G | 0.200 | 0.041 | 0.006 | 1.46E-12 | UHRF1 |
| rs17175860 | 19 | 7235146 | G | A | 0.192 | 0.045 | 0.006 | 2.55E-14 | INSR |
| rs2115107 | 19 | 7968168 | A | G | 0.392 | 0.038 | 0.005 | 8.45E-16 | LRRC8E,MAP2K7 |
| rs11666603 | 19 | 12496934 | C | T | 0.744 | 0.033 | 0.006 | 1.71E-09 | RPS29P23,ZNF799 |
| rs3111316 | 19 | 13038415 | A | G | 0.588 | 0.044 | 0.005 | 3.43E-21 | FARSA |
| rs10404726 | 19 | 18834514 | C | T | 0.531 | 0.028 | 0.005 | 2.73E-09 | CRTC1 |
| rs58542926 | 19 | 19379549 | T | C | 0.076 | 0.089 | 0.009 | 4.67E-25 | TM6SF2 |
| rs4805681 | 19 | 31835516 | C | T | 0.610 | 0.027 | 0.005 | 1.32E-08 | TSHZ3 |
| rs10406327 | 19 | 33890838 | C | G | 0.525 | 0.037 | 0.005 | 8.75E-16 | PEPD |
| rs429358 | 19 | 45411941 | T | C | 0.855 | 0.073 | 0.007 | 3.98E-28 | APOE |
| rs10407429 | 19 | 46157237 | G | A | 0.579 | 0.054 | 0.005 | 4.79E-31 | RN7SL836P,GIPR |
| rs2238689 | 19 | 46178661 | C | T | 0.418 | 0.039 | 0.005 | 5.40E-09 | GIPR |
| rs11667244 | 19 | 47580185 | G | A | 0.702 | 0.035 | 0.005 | 2.17E-12 | ZC3H4 |
| rs6515236 | 20 | 22435749 | A | C | 0.751 | 0.050 | 0.009 | 3.34E-08 | LOC105372562 (FOXA2) |
| rs2268078 | 20 | 32596704 | A | G | 0.655 | 0.039 | 0.005 | 2.14E-15 | RALY |
| rs17265513 | 20 | 39832628 | C | T | 0.198 | 0.033 | 0.006 | 1.13E-08 | ZHX3 |
| rs419842 | 20 | 42310811 | T | A | 0.821 | 0.041 | 0.006 | 7.17E-11 | MYBL2 |
| rs12625671 | 20 | 42994812 | C | T | 0.114 | 0.065 | 0.007 | 6.74E-19 | HNF4A |
| rs1800961 | 20 | 43042364 | T | C | 0.035 | 0.166 | 0.017 | 2.30E-22 | HNF4A |
| rs6066138 | 20 | 45594711 | G | A | 0.725 | 0.045 | 0.005 | 1.58E-18 | EYA2 |
| rs867489 | 20 | 48833957 | C | T | 0.539 | 0.031 | 0.005 | 1.51E-11 | PELATON,CEBPB |
| rs2426439 | 20 | 50999627 | C | T | 0.632 | 0.037 | 0.005 | 2.08E-14 | LINC01524 |
| rs2252221 | 20 | 51621922 | G | A | 0.531 | 0.025 | 0.005 | 4.48E-08 | TSHZ2 |
| rs911300 | 20 | 57387262 | G | A | 0.542 | 0.035 | 0.005 | 5.19E-14 | MIR296,PIEZO1P2 |
| rs1815591 | 20 | 61277014 | A | T | 0.398 | 0.034 | 0.005 | 5.50E-12 | SLCO4A1 |
| rs4809369 | 20 | 62470785 | G | A | 0.555 | 0.034 | 0.005 | 4.27E-13 | C20orf181,ZBTB46 |
| rs75756987 | 21 | 47767295 | G | C | 0.888 | 0.044 | 0.007 | 1.61E-09 | PCNT |
| rs75401573 | 22 | 29805444 | C | T | 0.924 | 0.051 | 0.009 | 6.47E-09 | RFPL4AP6,AP1B1 |
| rs5753043 | 22 | 30588041 | C | A | 0.898 | 0.061 | 0.008 | 3.76E-14 | LIF-AS1,HORMAD2 |
| rs117001013 | 22 | 32348841 | C | T | 0.919 | 0.047 | 0.008 | 1.44E-08 | YWHAH |
| rs133015 | 22 | 38572526 | C | G | 0.560 | 0.029 | 0.005 | 5.18E-10 | PLA2G6 |
| rs5751061 | 22 | 41593873 | G | T | 0.614 | 0.026 | 0.005 | 3.15E-08 | EP300-AS1,L3MBTL2 |
| rs3747207 | 22 | 44324855 | A | G | 0.221 | 0.047 | 0.006 | 9.26E-18 | PNPLA3 |
| rs5771069 | 22 | 50435480 | G | A | 0.505 | 0.033 | 0.005 | 1.78E-12 | IL17REL |
| rs112915006 | 22 | 50604696 | G | A | 0.050 | 0.092 | 0.015 | 7.50E-10 | NA |

chr, chromosome; pos, position; A1, effect allele; A2, other allele; eaf, effect allele frequency

**Supplementary Appendix 6. Quality assessment of each article**

1. Are the effect and other alleles coded in the same direction for the exposure and outcome (variable harmonization)?

2. Are the samples used to identify the genetic IVs for the risk factor and outcome drawn from the same ethnic population?

3. Were the two samples independent? (2SMR)

4. Assumption 1-Is there sufficient evidence that the genetic variants are robustly associated with the risk factor of interest?

5. Assumption 2-Are the genetic variants associated with potential confounders? Do the authors present this relationship?

6. Assumption 3-Is there any way for the genetic variants to affect the outcome through alternative pathways (horizontal pleiotropy)?

7. Was the analysis restricted to independent variants (that is, pruned of SNPs in linkage disequilibrium) or did the analysis allow for the correlation between variants?

8. Do the authors manually pick and choose which SNPs go into the instrument to tackle pleiotropy? If so, is the approach and justification clear?

9. Do the authors provide sensitivity analyses such as MR Egger, weighted median, and mode Mendelian randomisation, or use negative control populations?

10. Do the authors provide the data that they used (especially for Mendelian randomisation analyses conducted at the summary level) in a supplement to allow researchers to reproduce their findings?

**Supplementary Appendix 7. Quality assessment of the 8 articles article**

|  | (8) | (9) | (10) | (11) | (12) | (13) | (14) | (15) |
| --- | --- | --- | --- | --- | --- | --- | --- | --- |
| Q1 | ✔ | ✔ | ✔ | ✔ | ✔ | ✔ | ✔ | ✔ |
| Q2 | ✔ | ✔ | ✔ | ✔ | ✔ | ✔ | ✔ | ✔ |
| Q3 | ✔ | ✔ | ✔ | ✔ | ✔ | ✔ | ✔ | ✔ |
| Q4 | ✔ | ✔ | ✔ | ✔ | ✔ | ✔ | ✔ | ✔ |
| Q5 | ✔ | ✔ | ✔ | ✔ | ✔ | ✔ | ✔ | ✔ |
| Q6 | ✔ | ✔ | ✔ | ✔ | ✔ | ✔ | ✔ | ✔ |
| Q7 | ✔ | ✔ | ✔ | ✔ | ✔ | ✔ | ✔ | ✔ |
| Q8 | ✔ | ✔ | ✔ | ✔ | ✔ | ✔ | ✔ | ✔ |
| Q9 | ✔ | ✔ | ✔ | ✔ | ✔ | ✔ | ✔ | ✔ |
| Q10 | ✔ | ✔ | ✘ | ✔ | ✔ | ✔ | ✔ | ✔ |

**Supplementary Appendix 8. Result of MR analysis between type 2 diabetes mellitus and Alzheimer’s disease (IGAP dataset)**

| Method | **nSNP** | **OR** | **95% CI** | **p-value** | **MR Egger intercept** | **I^2^** |
| --- | --- | --- | --- | --- | --- | --- |
| MR Egger | 440 | 0.885 | 0.725-1.079 | 0.228 | 0.437 | 0.00% |
| Weighted median | 440 | 0.987 | 0.917-1.061 | 0.716 |  |  |
| Inverse variance weighted | 440 | 0.947 | 0.856-1.048 | 0.291 |  |  |
| Simple mode | 440 | 1.106 | 0.924-1.322 | 0.272 |  |  |
| Weighted mode | 440 | 0.991 | 0.914-1.075 | 0.823 |  |  |

SNP, single nucleotide polymorphism; OR, odds ratio; CI: confidence interval

**Supplementary Appendix 9. Result of MR analysis between type 2 diabetes mellitus (T2DM) and Alzheimer’s disease (EDAB dataset)**

| Method | **nSNP** | **OR** | **95% CI** | **p-value** | **MR Egger intercept** | **I^2^** |
| --- | --- | --- | --- | --- | --- | --- |
| MR Egger | 490 | 1.009 | 0.963-1.057 | 0.721 | 0.424 | 27.14% |
| Weighted median | 490 | 1.013 | 0.979-1.048 | 0.453 |  |  |
| Inverse variance weighted | 490 | 0.992 | 0.969-1.016 | 0.522 |  |  |
| Simple mode | 490 | 1.075 | 0.989-1.17 | 0.091 |  |  |
| Weighted mode | 490 | 1.029 | 0.989-1.070 | 0.161 |  |  |

SNP, single nucleotide polymorphism; OR, odds ratio; CI: confidence interval

**Supplementary Appendix 10. Result of MR analysis between type 2 diabetes mellitus (T2DM) and Alzheimer’s disease (UKB dataset)**

| Method | **nSNP** | **OR** | **95% CI** | **p-value** | **MR Egger intercept** | **I^2^** |
| --- | --- | --- | --- | --- | --- | --- |
| MR Egger | 492 | 1.00 | 0.999-1.000 | 0.698 | 0.629 | 0.00% |
| Weighted median | 492 | 1.0002 | 1.000-1.001 | 0.296 |  |  |
| Inverse variance weighted | 492 | 1.0000 | 1.000-1.000 | 0.955 |  |  |
| Simple mode | 492 | 0.9999 | 0.999-1.001 | 0.788 |  |  |
| Weighted mode | 492 | 1.0001 | 1.000-1.001 | 0.551 |  |  |

SNP, single nucleotide polymorphism; OR, odds ratio; CI: confidence interval

**Supplementary Appendix 11. PRISMA checklist**

| **Section and Topic** | **Item #** | **Checklist item** | **Location where item is reported** |
| --- | --- | --- | --- |
| **TITLE** | | |  |
| Title | 1 | Identify the report as a systematic review. | Page 1 |
| **ABSTRACT** | | |  |
| Abstract | 2 | See the PRISMA 2020 for Abstracts checklist. | Page 2-3 |
| **INTRODUCTION** | | |  |
| Rationale | 3 | Describe the rationale for the review in the context of existing knowledge. | Page 4 |
| Objectives | 4 | Provide an explicit statement of the objective(s) or question(s) the review addresses. | Page 5 |
| **METHODS** | | |  |
| Eligibility criteria | 5 | Specify the inclusion and exclusion criteria for the review and how studies were grouped for the syntheses. | Page 6 |
| Information sources | 6 | Specify all databases, registers, websites, organisations, reference lists and other sources searched or consulted to identify studies. Specify the date when each source was last searched or consulted. | Page 5 |
| Search strategy | 7 | Present the full search strategies for all databases, registers and websites, including any filters and limits used. | Page 6 |
| Selection process | 8 | Specify the methods used to decide whether a study met the inclusion criteria of the review, including how many reviewers screened each record and each report retrieved, whether they worked independently, and if applicable, details of automation tools used in the process. | Page 6 |
| Data collection process | 9 | Specify the methods used to collect data from reports, including how many reviewers collected data from each report, whether they worked independently, any processes for obtaining or confirming data from study investigators, and if applicable, details of automation tools used in the process. | Page 6 |
| Data items | 10a | List and define all outcomes for which data were sought. Specify whether all results that were compatible with each outcome domain in each study were sought (e.g. for all measures, time points, analyses), and if not, the methods used to decide which results to collect. | Page 6 |
|  | 10b | List and define all other variables for which data were sought (e.g. participant and intervention characteristics, funding sources). Describe any assumptions made about any missing or unclear information. | Page 8 |
| Study risk of bias assessment | 11 | Specify the methods used to assess risk of bias in the included studies, including details of the tool(s) used, how many reviewers assessed each study and whether they worked independently, and if applicable, details of automation tools used in the process. | Page 7 |
| Effect measures | 12 | Specify for each outcome the effect measure(s) (e.g. risk ratio, mean difference) used in the synthesis or presentation of results. | Page 6 |
| Synthesis methods | 13a | Describe the processes used to decide which studies were eligible for each synthesis (e.g. tabulating the study intervention characteristics and comparing against the planned groups for each synthesis (item #5)). | Page 6 |
|  | 13b | Describe any methods required to prepare the data for presentation or synthesis, such as handling of missing summary statistics, or data conversions. | Page 6 |
|  | 13c | Describe any methods used to tabulate or visually display results of individual studies and syntheses. | Page 6 |
|  | 13d | Describe any methods used to synthesize results and provide a rationale for the choice(s). If meta-analysis was performed, describe the model(s), method(s) to identify the presence and extent of statistical heterogeneity, and software package(s) used. | Page 7 |
|  | 13e | Describe any methods used to explore possible causes of heterogeneity among study results (e.g. subgroup analysis, meta-regression). | Page 7 |
|  | 13f | Describe any sensitivity analyses conducted to assess robustness of the synthesized results. | / |
| Reporting bias assessment | 14 | Describe any methods used to assess risk of bias due to missing results in a synthesis (arising from reporting biases). | / |
| Certainty assessment | 15 | Describe any methods used to assess certainty (or confidence) in the body of evidence for an outcome. | / |
| **RESULTS** | | |  |
| Study selection | 16a | Describe the results of the search and selection process, from the number of records identified in the search to the number of studies included in the review, ideally using a flow diagram. | Page 12 |
|  | 16b | Cite studies that might appear to meet the inclusion criteria, but which were excluded, and explain why they were excluded. | Page 12 |
| Study characteristics | 17 | Cite each included study and present its characteristics. | Page 12 |
| Risk of bias in studies | 18 | Present assessments of risk of bias for each included study. | Page 12 |
| Results of individual studies | 19 | For all outcomes, present, for each study: (a) summary statistics for each group (where appropriate) and (b) an effect estimate and its precision (e.g. confidence/credible interval), ideally using structured tables or plots. | Page 25 |
| Results of syntheses | 20a | For each synthesis, briefly summarise the characteristics and risk of bias among contributing studies. | Supplementary Appendix 6 |
|  | 20b | Present results of all statistical syntheses conducted. If meta-analysis was done, present for each the summary estimate and its precision (e.g. confidence/credible interval) and measures of statistical heterogeneity. If comparing groups, describe the direction of the effect. | Supplementary Appendix 2 |
|  | 20c | Present results of all investigations of possible causes of heterogeneity among study results. | Page 12 |
|  | 20d | Present results of all sensitivity analyses conducted to assess the robustness of the synthesized results. | / |
| Reporting biases | 21 | Present assessments of risk of bias due to missing results (arising from reporting biases) for each synthesis assessed. | / |
| Certainty of evidence | 22 | Present assessments of certainty (or confidence) in the body of evidence for each outcome assessed. | / |
| **DISCUSSION** | | |  |
| Discussion | 23a | Provide a general interpretation of the results in the context of other evidence. | Page 14 |
|  | 23b | Discuss any limitations of the evidence included in the review. | Page 17 |
|  | 23c | Discuss any limitations of the review processes used. | / |
|  | 23d | Discuss implications of the results for practice, policy, and future research. | Page 16 |
| **OTHER INFORMATION** | | |  |
| Registration and protocol | 24a | Provide registration information for the review, including register name and registration number, or state that the review was not registered. | Page 5 |
|  | 24b | Indicate where the review protocol can be accessed, or state that a protocol was not prepared. | Page 5 |
|  | 24c | Describe and explain any amendments to information provided at registration or in the protocol. | Page 5 |
| Support | 25 | Describe sources of financial or non-financial support for the review, and the role of the funders or sponsors in the review. | Page 18 |
| Competing interests | 26 | Declare any competing interests of review authors. | Page 18 |
| Availability of data, code and other materials | 27 | Report which of the following are publicly available and where they can be found: template data collection forms; data extracted from included studies; data used for all analyses; analytic code; any other materials used in the review. | Page 12 |

*From:* Page MJ, McKenzie JE, Bossuyt PM, Boutron I, Hoffmann TC, Mulrow CD, et al. The PRISMA 2020 statement: an updated guideline for reporting systematic reviews. BMJ 2021;372:n71. doi: 10.1136/bmj.n71. This work is licensed under CC BY 4.0. To view a copy of this license, visit <https://creativecommons.org/licenses/by/4.0/>

**Supplementary Appendix 12. STROBE-MR checklist of recommended items to address in reports of Mendelian randomization studies**

| **Item No.** | **Section** | **Checklist item** | **Page No.** | **Relevant text from manuscript** |
| --- | --- | --- | --- | --- |
| 1 | **TITLE and ABSTRACT** | Indicate Mendelian randomization (MR) as the study’s design in the title and/or the abstract if that is a main purpose of the study | 1 | Title and Abstract |
|  | **INTRODUCTION** |  |  |  |
| 2 | **Background** | Explain the scientific background and rationale for the reported study. What is the exposure? Is a potential causal relationship between exposure and outcome plausible? Justify why MR is a helpful method to address the study question | 4 | Introduction |
| 3 | **Objectives** | State specific objectives clearly, including pre-specified causal hypotheses (if any). State that MR is a method that, under specific assumptions, intends to estimate causal effects | 5 | Introduction |
|  | **METHODS** |  |  |  |
| 4 | **Study design and data sources** | Present key elements of the study design early in the article. Consider including a table listing sources of data for all phases of the study. For each data source contributing to the analysis, describe the following: |  |  |
|  | a) | Setting: Describe the study design and the underlying population, if possible. Describe the setting, locations, and relevant dates, including periods of recruitment, exposure, follow-up, and data collection, when available. | 7 | Methods- Two-sample MR analysis |
|  | b) | Participants: Give the eligibility criteria, and the sources and methods of selection of participants. Report the sample size, and whether any power or sample size calculations were carried out prior to the main analysis | 5 | Methods- Systematic Review and Meta-MR |
|  | c) | Describe measurement, quality control and selection of genetic variants | 8 | Methods- Instrumental variables selection |
|  | d) | For each exposure, outcome, and other relevant variables, describe methods of assessment and diagnostic criteria for diseases | S3 Appendix | Methods- Information of instrumental variables consortiums |
|  | e) | Provide details of ethics committee approval and participant informed consent, if relevant | 10 | Methods- Outcome Genetic Consortia Data |
| 5 | **Assumptions** | Explicitly state the three core IV assumptions for the main analysis (relevance, independence and exclusion restriction) as well assumptions for any additional or sensitivity analysis | 11  24 | Methods- Statistical Methods and Sensitivity Analyses  Figure 2 |
| 6 | **Statistical methods: main analysis** | Describe statistical methods and statistics used |  |  |
|  | a) | Describe how quantitative variables were handled in the analyses (i.e., scale, units, model) | 11 | Methods- Statistical Methods and Sensitivity Analyses |
|  | b) | Describe how genetic variants were handled in the analyses and, if applicable, how their weights were selected | 11 | Methods- Statistical Methods and Sensitivity Analyses |
|  | c) | Describe the MR estimator (e.g. two-stage least squares, Wald ratio) and related statistics. Detail the included covariates and, in case of two-sample MR, whether the same covariate set was used for adjustment in the two samples | 11 | Methods- Statistical Methods and Sensitivity Analyses |
|  | d) | Explain how missing data were addressed | 8 | Methods- Two-sample MR analysis |
|  | e) | If applicable, indicate how multiple testing was addressed | / |  |
| 7 | **Assessment of assumptions** | Describe any methods or prior knowledge used to assess the assumptions or justify their validity | 9 | Methods- Instrumental variables selection |
| 8 | **Sensitivity analyses and additional analyses** | Describe any sensitivity analyses or additional analyses performed (e.g. comparison of effect estimates from different approaches, independent replication, bias analytic techniques, validation of instruments, simulations) | 9 | Methods- Instrumental variables selection |
| 9 | **Software and pre-registration** |  |  |  |
|  | a) | Name statistical software and package(s), including version and settings used | 12 | Methods- Statistical Methods and Sensitivity Analyses |
|  | b) | State whether the study protocol and details were pre-registered (as well as when and where) | / | / |
|  | **RESULTS** |  |  |  |
| 10 | **Descriptive data** |  |  |  |
|  | a) | Report the numbers of individuals at each stage of included studies and reasons for exclusion. Consider use of a flow diagram | 12  23 | Findings- Systematic Review and Meta-MR  Figure 1 |
|  | b) | Report summary statistics for phenotypic exposure(s), outcome(s), and other relevant variables (e.g. means, SDs, proportions) | / | Supplementary Appendix 4 |
|  | c) | If the data sources include meta-analyses of previous studies, provide the assessments of heterogeneity across these studies | 6 | Methods- Systematic Review and Meta-MR |
|  | d) | For two-sample MR:  i.  Provide justification of the similarity of the genetic variant-exposure associations between the exposure and outcome samples  ii.  Provide information on the number of individuals who overlap between the exposure and outcome studies | / | Supplementary Appendix 2 |
| 11 | **Main results** |  |  |  |
|  | a) | Report the associations between genetic variant and exposure, and between genetic variant and outcome, preferably on an interpretable scale | 13 | Findings- Two-Sample MR analysis |
|  | b) | Report MR estimates of the relationship between exposure and outcome, and the measures of uncertainty from the MR analysis, on an interpretable scale, such as odds ratio or relative risk per SD difference | 13 | Findings- Two-Sample MR analysis |
|  | c) | If relevant, consider translating estimates of relative risk into absolute risk for a meaningful time period | / | / |
|  | d) | Consider plots to visualize results (e.g. forest plot, scatterplot of associations between genetic variants and outcome versus between genetic variants and exposure) | 26-28 | Figure 4-6 |
| 12 | **Assessment of assumptions** |  |  |  |
|  | a) | Report the assessment of the validity of the assumptions | 8 | Methods- Two-Sample MR analysis- Instrumental variables selection |
|  | b) | Report any additional statistics (e.g., assessments of heterogeneity across genetic variants, such as *I^2^*, Q statistic or E-value) | 13-14 | Findings- Two-Sample MR analysis |
| 13 | **Sensitivity analyses and additional analyses** |  |  |  |
|  | a) | Report any sensitivity analyses to assess the robustness of the main results to violations of the assumptions | 13-14 | Findings- Two-Sample MR analysis |
|  | b) | Report results from other sensitivity analyses or additional analyses | / | / |
|  | c) | Report any assessment of direction of causal relationship (e.g., bidirectional MR) | / | / |
|  | d) | When relevant, report and compare with estimates from non-MR analyses | / | / |
|  | e) | Consider additional plots to visualize results (e.g., leave-one-out analyses) | / | / |
|  | **DISCUSSION** |  |  |  |
| 14 | **Key results** | Summarize key results with reference to study objectives | 14 | Discussion |
| 15 | **Limitations** | Discuss limitations of the study, taking into account the validity of the IV assumptions, other sources of potential bias, and imprecision. Discuss both direction and magnitude of any potential bias and any efforts to address them | 17 | Discussion- Strengths and limitations |
| 16 | **Interpretation** |  |  |  |
|  | a) | Meaning: Give a cautious overall interpretation of results in the context of their limitations and in comparison with other studies | 14 | Discussion |
|  | b) | Mechanism: Discuss underlying biological mechanisms that could drive a potential causal relationship between the investigated exposure and the outcome, and whether the gene-environment equivalence assumption is reasonable. Use causal language carefully, clarifying that IV estimates may provide causal effects only under certain assumptions | 14 | Discussion |
|  | c) | Clinical relevance: Discuss whether the results have clinical or public policy relevance, and to what extent they inform effect sizes of possible interventions | 16 | Discussion-Clinical relevance |
| 17 | **Generalizability** | Discuss the generalizability of the study results (a) to other populations, (b) across other exposure periods/timings, and (c) across other levels of exposure | 14 | Discussion |
|  | **OTHER INFORMATION** |  |  |  |
| 18 | **Funding** | Describe sources of funding and the role of funders in the present study and, if applicable, sources of funding for the databases and original study or studies on which the present study is based | 18 | Acknowledgements |
| 19 | **Data and data sharing** | Provide the data used to perform all analyses or report where and how the data can be accessed, and reference these sources in the article. Provide the statistical code needed to reproduce the results in the article, or report whether the code is publicly accessible and if so, where | 12 | Methods- Statistical Methods and Sensitivity Analyses |
| 20 | **Conflicts of Interest** | All authors should declare all potential conflicts of interest | 18 | Acknowledgements |

This checklist is copyrighted by the Equator Network under the Creative Commons Attribution 3.0 Unported (CC BY 3.0) license.

1.	Skrivankova VW, Richmond RC, Woolf BAR, Yarmolinsky J, Davies NM, Swanson SA, et al. Strengthening the Reporting of Observational Studies in Epidemiology using Mendelian Randomization (STROBE-MR) Statement. JAMA. 2021;under review.

2.	Skrivankova VW, Richmond RC, Woolf BAR, Davies NM, Swanson SA, VanderWeele TJ, et al. Strengthening the Reporting of Observational Studies in Epidemiology using Mendelian Randomisation (STROBE-MR): Explanation and Elaboration. BMJ. 2021;375:n2233.

1.	Scott RA, Scott LJ, Mägi R, Marullo L, Gaulton KJ, Kaakinen M, et al. An Expanded Genome-Wide Association Study of Type 2 Diabetes in Europeans. Diabetes. 2017;66(11):2888-902.

2.	Mahajan A, Wessel J, Willems SM, Zhao W, Robertson NR, Chu AY, et al. Refining the accuracy of validated target identification through coding variant fine-mapping in type 2 diabetes. Nat Genet. 2018;50(4):559-71.

3.	Fuchsberger C, Flannick J, Teslovich TM, Mahajan A, Agarwala V, Gaulton KJ, et al. The genetic architecture of type 2 diabetes. Nature. 2016;536(7614):41-7.

4.	Sudlow C, Gallacher J, Allen N, Beral V, Burton P, Danesh J, et al. UK biobank: an open access resource for identifying the causes of a wide range of complex diseases of middle and old age. PLoS Med. 2015;12(3):e1001779.

5.	Cook JP, Morris AP. Multi-ethnic genome-wide association study identifies novel locus for type 2 diabetes susceptibility. Eur J Hum Genet. 2016;24(8):1175-80.

6.	Estrada K, Aukrust I, Bjørkhaug L, Burtt NP, Mercader JM, García-Ortiz H, et al. Association of a low-frequency variant in HNF1A with type 2 diabetes in a Latino population. Jama. 2014;311(22):2305-14.

7.	Vujkovic M, Keaton JM, Lynch JA, Miller DR, Zhou J, Tcheandjieu C, et al. Discovery of 318 new risk loci for type 2 diabetes and related vascular outcomes among 1.4 million participants in a multi-ancestry meta-analysis. Nat Genet. 2020;52(7):680-91.

8.	Østergaard SD, Mukherjee S, Sharp SJ, Proitsi P, Lotta LA, Day F, et al. Associations between Potentially Modifiable Risk Factors and Alzheimer Disease: A Mendelian Randomization Study. PLoS Med. 2015;12(6):e1001841; discussion e.

9.	Andrews SJ, Fulton-Howard B, O'Reilly P, Marcora E, Goate AM. Causal Associations Between Modifiable Risk Factors and the Alzheimer's Phenome. Ann Neurol. 2021;89(1):54-65.

10.	Meng L, Wang Z, Ji HF, Shen L. Causal association evaluation of diabetes with Alzheimer's disease and genetic analysis of antidiabetic drugs against Alzheimer's disease. Cell Biosci. 2022;12(1):28.

11.	Luo J, Thomassen JQ, Bellenguez C, Grenier-Boley B, de Rojas I, Castillo A, et al. Genetic Associations Between Modifiable Risk Factors and Alzheimer Disease. JAMA Netw Open. 2023;6(5):e2313734.

12.	Pan Y, Chen W, Yan H, Wang M, Xiang X. Glycemic traits and Alzheimer's disease: a Mendelian randomization study. Aging (Albany NY). 2020;12(22):22688-99.

13.	Dybjer E, Kumar A, Nägga K, Engström G, Mattsson-Carlgren N, Nilsson PM, et al. Polygenic risk of type 2 diabetes is associated with incident vascular dementia: a prospective cohort study. Brain Commun. 2023;5(2):fcad054.

14.	Garfield V, Farmaki AE, Fatemifar G, Eastwood SV, Mathur R, Rentsch CT, et al. Relationship Between Glycemia and Cognitive Function, Structural Brain Outcomes, and Dementia: A Mendelian Randomization Study in the UK Biobank. Diabetes. 2021;70(10):2313-21.

15.	Thomassen JQ, Tolstrup JS, Benn M, Frikke-Schmidt R. Type-2 diabetes and risk of dementia: observational and Mendelian randomisation studies in 1 million individuals. Epidemiol Psychiatr Sci. 2020;29:e118.
